# Potential health and economic impact of paediatric vaccination using next generation influenza vaccines in Kenya: a modelling study

**DOI:** 10.1101/2022.08.26.22279262

**Authors:** Naomi R Waterlow, Sreejith Radhakrishnan, Jeanette Dawa, Edwin van Leeuwen, Philipp Lambach, Joseph Bresee, Marie Mazur, Rosalind M Eggo, Mark Jit

## Abstract

**Background:** Influenza is a major year-round cause of respiratory illness in Kenya, particularly in children under 5. Current influenza vaccines result in short-term, strain-specific immunity and were found in a previous study not to be cost-effective in Kenya. However, next generation vaccines are in development that may have a greater impact and cost-effectiveness profile.

**Methods:** We expanded a model previously used to evaluate cost-effectiveness of seasonal influenza vaccines in Kenya to include next generation vaccines by allowing for enhanced vaccine characteristics and multi-annual immunity. We specifically examined vaccinating children under 5 years of age with improved vaccines, evaluating vaccines with combinations of increased vaccine effectiveness, cross protection between strains (breadth) and duration of immunity. We evaluated cost-effectiveness using incremental cost-effectiveness ratios (ICERs) and incremental net monetary benefits (INMBs) for a range of values for the willingness-to-pay (WTP) per DALY averted. Finally, we estimated threshold per-dose vaccine prices at which vaccination becomes cost-effective.

**Results:** Next generation vaccines can be cost-effective, dependent on the vaccine characteristics and assumed WTP thresholds. Universal vaccines (assumed to provide long-term and broad immunity) are most cost-effective in Kenya across three of four WTP thresholds evaluated, with the lowest median value of ICER per DALY averted ($263, 95% Credible Interval (CrI): $-1698, $1061) and the highest median INMBs. At a WTP of $623, universal vaccines are cost-effective at or below a median price of $5.16 per dose (95% CrI: $0.94, $18.57). We also show that the assumed mechanism underlying infection-derived immunity strongly impacts vaccine outcomes.

**Conclusion:** This evaluation provides evidence for country-level decision makers about future next generation vaccine introduction, as well as global research funders about the potential market for these vaccines. Next generation vaccines may offer a cost-effective intervention to reduce influenza burden in low-income countries with year-round seasonality like Kenya.

## Background

Influenza is a major cause of respiratory illness in Kenya, particularly in children under 5 years old (1,2). Current influenza vaccines result in short-term, strain-specific immunity (3) which is particularly problematic in tropical and subtropical settings where multiple peaks and identifiable year-round activity make it challenging to decide if, who and when to vaccinate, as well as which formulation (northern or southern hemisphere) to use (4–7). Existing vaccines have been evaluated for cost-effectiveness in Kenya, looking at the potential impact of vaccinations in 2010 to 2018 (2). This analysis showed that vaccinating children in Kenya with currently available vaccines was not cost-effective, given current willingness-to-pay thresholds (2). Barriers to cost-effectiveness of influenza vaccination include inconsistent seasonality (with high burden across the year in some years), multiple subtypes of influenza, varying vaccine effectiveness depending on match to circulating influenza strains and the need for annual revaccination(8).

Many of these obstacles could be addressed by next generation vaccines on the near horizon, with 18 vaccines in clinical trials (10 in phase I, 6 in phase II and 2 in phase III trials), and over 100 in preclinical trials (9,10). Newer technologies are being trialled, for example mRNA vaccines and self-assembling nano-particles, and many of these vaccines aim to overcome the immunodominance of the haemagglutinin (HA) head, instead focusing on more conserved proteins across influenza strains and often aiming to stimulate a T-cell response (9).

The World Health Organisation (WHO) Preferred Product Characteristics (PPC) (11) describes next generation influenza vaccines in two categories: improved vaccines, which have increased vaccine efficacy (VE) or strain cross-protection (breadth) and which generate immune protection lasting at least a year; and universal vaccines, which have increased efficacy against influenza A phylogenetic HA group viruses and which generate immune protection lasting at least 5 years.These descriptions are based on the likelihood of development in the near to mid future. The US National Institute of Allergy and Infectious Diseases (NIAID) uses similar but slightly varying definitions (12). Such next generation vaccines may hold promising benefits for countries like Kenya, but their potential population impact and cost-effectiveness have yet to be evaluated. Such evaluations could inform country-level decision makers about potential future vaccine introduction, as well as global research funders about the potential market for these vaccines.

Mathematical models are ideal tools for evaluating their cost-effectiveness, as they allow analysis and comparison of potential hypothetical interventions and strategies. Specifically, transmission dynamic models have the additional advantage of including both direct and indirect benefits of vaccination. This allows evaluation of optimal control strategies including coverage and timing of vaccination campaigns and vaccine characteristics, such as subtype broadness vs efficacy considerations.

We expand the model previously used to evaluate cost-effectiveness of current influenza seasonal vaccines in Kenya to evaluate the cost-effectiveness of next generation vaccines.

## Methods

### Overview

We utilise a transmission model from Baguelin *et al*. (2013) (13) that was fitted to Kenya severe acute respiratory illness (SARI) data from 2010 - 2018 by Dawa *et al*. (2013) (2) and extend it to include next generation influenza vaccines with longer durations of immunity, higher efficacy and/or broader sub-type cross-protection (Figure 1). Code is available at https://github.com/NaomiWaterlow/NextGenFlu_Kenya

**Figure 1.**
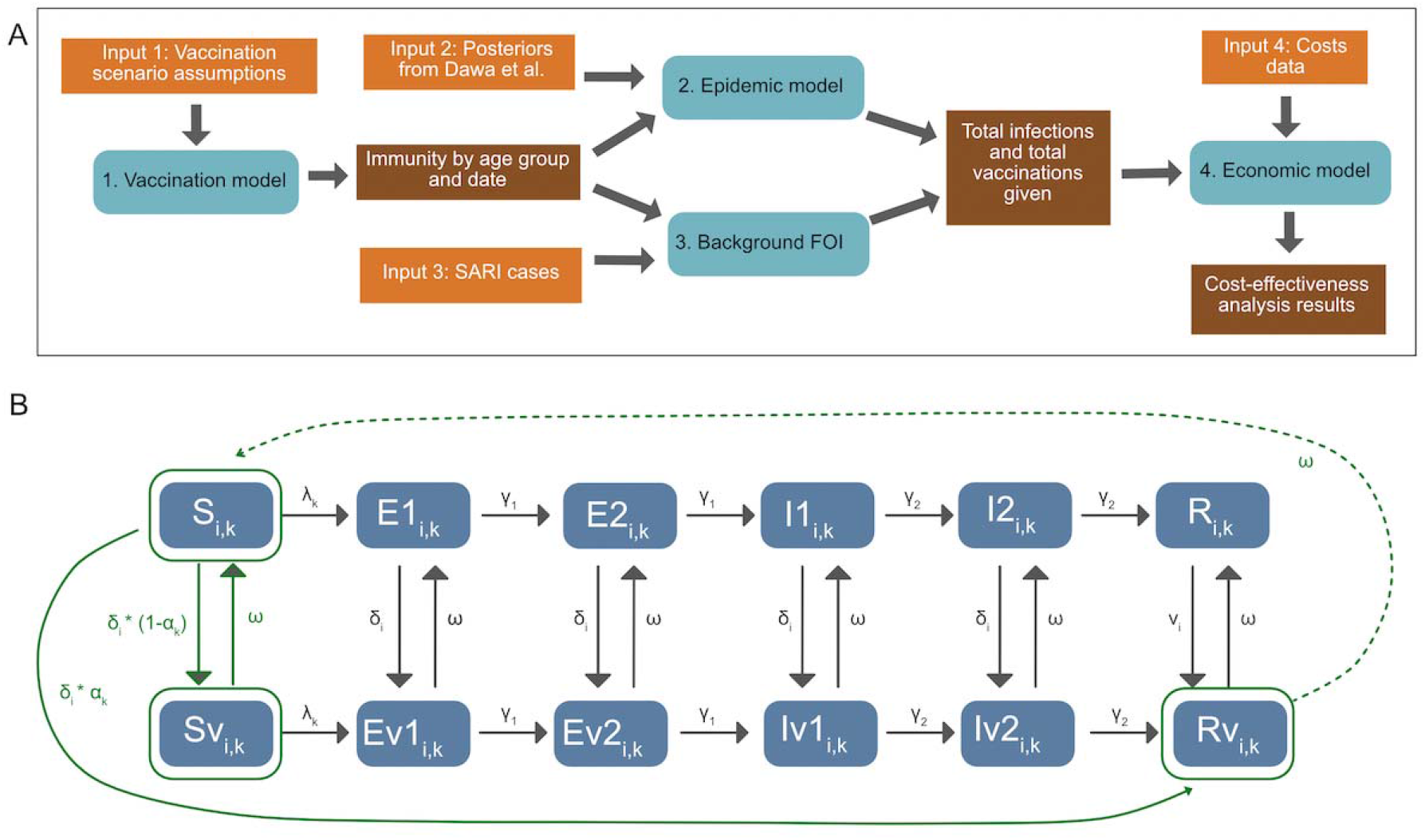
Modelling overview: A) Methods overview, depicting inputs, models and outputs. B) Model diagram, including both the epidemic and the vaccination model. Elements in solid green are included in both models. Transitions in grey are included only in the epidemic model, and transitions in dotted green are included only in the vaccination model. States are: Susceptible (S), Exposed (E), Infectious (I) and Recovered (R), and their vaccinated counterparts (Sv, Ev1, Ev2, Iv1, Iv2, Rv). v denotes the vaccinated equivalent of the compartments. See Table S3 for parameter details. is the rate of vaccination in age group, is the efficacy by subtype (), is vaccine derived immunity waning. The model is run separately for each subtype. For the epidemic model, in both vaccinated and unvaccinated compartments, susceptibles who are infected with the viral subtype enter the first Exposed (E) compartment. They then progress through the E and Infectious (I) compartments. After ceasing to be infectious they enter the R compartment, whereupon they cannot be re-infected during the same epidemic period. Both the E and I populations consist of two compartments, in order to get a gamma distributed waiting time. Each compartment is also subdivided by age (i).

### Model 1 - Vaccination model

The vaccination model (Figure 1B - green compartments) tracks the dynamics of vaccine-induced immunity for each virus subtype without considering prior infection or vaccination status. This is a conservative assumption, assuming vaccination status in the population is unknown and hence people are vaccinated independent of whether they were recently infected or vaccinated. At the time of vaccination the population can be in 1 of 3 compartments: Susceptible (*S*), Susceptible-vaccinated (*Sv*) and Recovered-vaccinated (*Rv*) (Figure 1A, green). Vaccination is assumed to be all-or-nothing, with a proportion defined by the efficacy for each subtype entering the *Rv* compartment where they are immune, and the inverse proportion entering the *Sv* compartment, where they are susceptible. Waning of vaccination from compartments *Sv* and *Rv* occurs exponentially at a rate, *ω*, determined by the duration of vaccine-induced immunity, returning the population to the *Sv* compartment.

We consider scenarios where vaccines have characteristics matching either currently available seasonal influenza vaccines, or next generation vaccines in line with WHO Preferred Product Characteristics (11) (input 1). In the first year, all 0-5 year olds are vaccinated across all vaccine scenarios. Following this, vaccination occurs every x years, calculated as a proportion of the age group, where x is the mean duration of vaccine derived immunity. We generate 5 vaccine scenario examples, corresponding to four categories of Preferred Product Characteristics: current seasonal vaccines, minimally improved vaccines, improved efficacy vaccines, improved breadth vaccines, and universal vaccines (Table 1). We consider vaccines to be either ‘matched’ or ‘mis-matched’ to circulating strains each season, and a different efficacy is given in these cases (see supplement for more details).

**Table 1:** Illustrative vaccine scenarios. “Mis-matched seasons” refers to the possibility that the vaccine is not well matched to a particular season’s influenza strain and therefore has reduced efficacy. Immunity duration is assumed to be exponential. All vaccines are given as a campaign, across March, April and May.

| Scenario name | Mis-matched seasons? | Efficacy (Matched/Mis-matched) | Immunity Duration | Coverage | Age-groups vaccinated |
| --- | --- | --- | --- | --- | --- |
| No Vaccine | - | - | - | - | - |
| Current Seasonal Vaccines | Yes | 70% / 40% | 6 months | 50% | All 0-5 |
| Improved vaccines (Minimal) | Yes | 70% / 40% | 1 year | 50% | All 0-5 |
| Improved Vaccines (Efficacy) | Yes | 90% / 70% | 2 years | 50% | All individuals aged 0-5 in the first year of vaccination, followed by age 0, 2 and 4 in subsequent years |
| Improved Vaccines (Breadth) | No | 70% / 70% | 3 years | 50% | All individuals aged 0-5 in the first year of vaccination, followed by age 0, 2 and 4 in subsequent years |
| Universal Vaccines | No | 90% / 90% | 5 years | 50% | All individuals aged 0-5 in the first year of vaccination,<br>followed by age 0<br>and 5 |

We also run sensitivity analyses where vaccination coverage is 75% for all vaccines, allowing for higher uptake upon new vaccine development (Supplement Section 9).

We run the model from 1 March 2010, where 1 March each year is considered the start of the southern hemisphere (SH) influenza season. The model runs with given inputs until 31 August, as we define 1 September as the start of the Northern Hemisphere (NH) influenza season. VE can differ between seasons to take account of vaccine matched or mis-matched strains. As in Dawa *et al*. (2020) we identify each season’s strain as matched or mis-matched to vaccination based on published VE data (Table S1) and assume that a VE >= 50% is a matched vaccine, and < 50% is a mis-matched vaccine. Following the NH season, the population size is updated (see Supplement section 1), and ageing of the population occurs, to allow for a build up of immunity in the relevant age groups. The model runs from 1 March 2010 to 28 February 2019. We model transmission of each influenza subtype separately (A(H1N1), A(H3N2), B) to allow different vaccine efficacies across subtypes. We assume all individuals are born susceptible to infection.

This vaccination model outputs the proportion of the population that is vaccinated, and of this the proportion that is immunised for each subtype every week over the modelled period.

### Model 2 - Epidemic Model

We model the 11 subtype-specific epidemic time periods that were identified and fitted in Dawa *et al*. (2020) (Figure 2A). As in Dawa *et al*. (2020) we define influenza epidemics to start at the first week of a time period consisting of “≥2 successive weeks where the proportion of subtype-specific test-positive cases was greater than the average weekly proportion during the entire study” (Figure 2A). Where an epidemic was previously defined to last less than 8 months, we follow it for the full 8 months to allow capturing the consequences of a slower epidemic progression as the result of vaccination. At the start of each epidemic the proportion of the population in the *S, Sv* and *Rv* compartments is taken from the output of the vaccination model, in the matching week and for the relevant virus subtype.Vaccine efficacy is split into NH and SH time frames as in the vaccination model. For each epidemic we run an independent transmission model (with structure of Figure 1B) with the estimated transmission rate, susceptibility for three age groups (<= 14, 15-49, 50+), initial number of infections and the probability of identifying an influenza-positive patient within the catchment population for 3 age groups (<1, 1-5, 6+) from Dawa *et al*. (2020). Influenza immunity is assumed to be leaky. Supplement section 2 contains the model equations, parameters and values.

**Figure 2:**
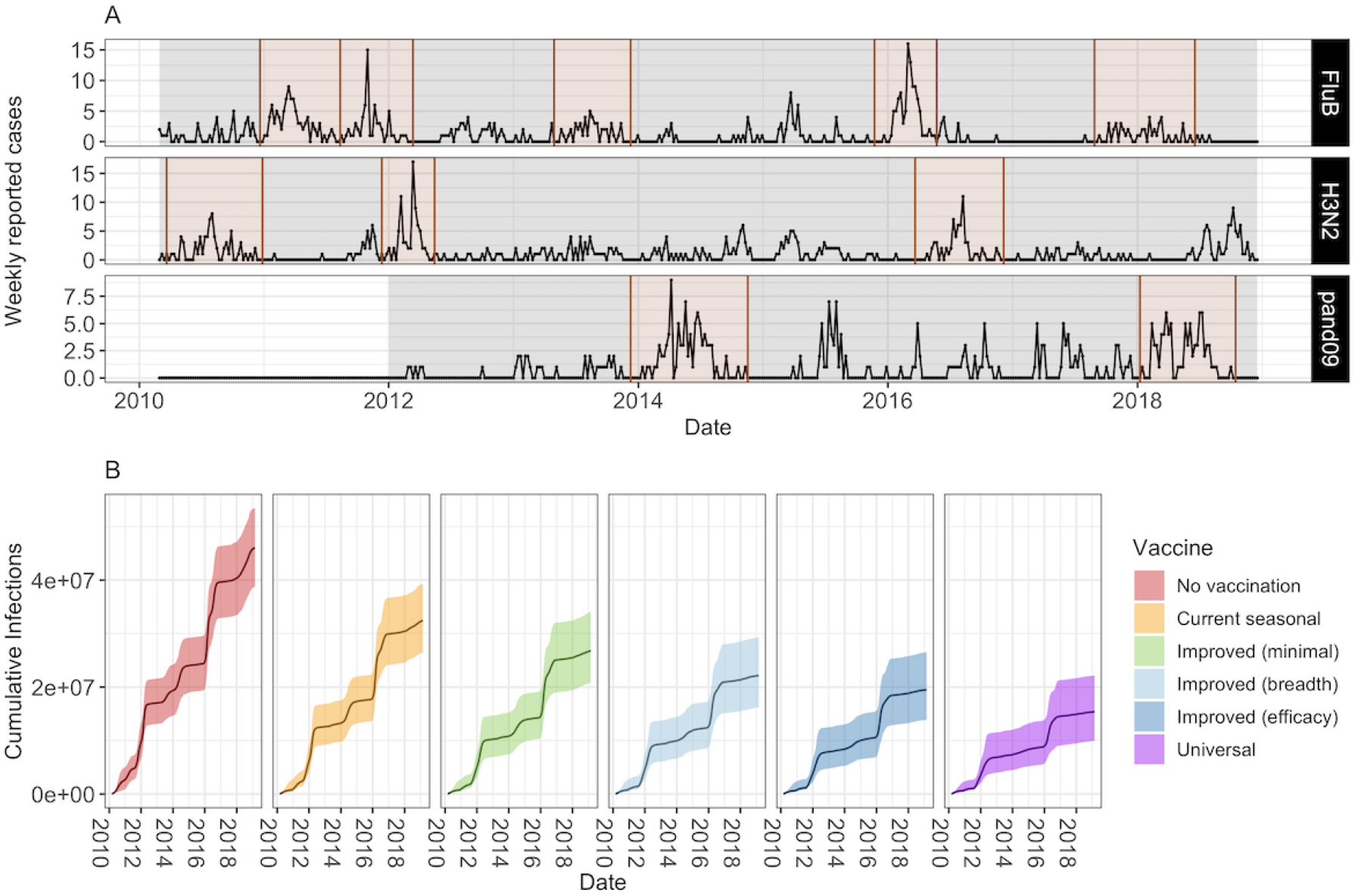
A) Number of weekly reported cases during epidemic and inter-epidemic time periods. Epidemic periods are highlighted in brown, and periods used to estimate the background force of infection are shown in grey. B) Model projections of cumulative number of infections (median and 95% CrI) by vaccine scenario.

For key transmission parameters (transmission rate, susceptibility, number of infections at the start of the season, number of imports and ascertainment rates), we use the estimated values by Dawa *et al*. 2020 for each of the 11 strain/subtype-specific peaks in influenza activity identified between 2010 - 2018 (input 2). The parameter values for each strain/subtype-specific peak are estimated independently, using the *fluEvidenceSynthesis* R package. We also use the same age groups (< 1, 1–5, 6–14, 15–19, 20–49 and ≥ 50 years old) contact patterns and population sizes. For more details see Dawa *et al*. (2020).

In our main analysis we assume that the previous season’s vaccination has no effect on the proportion of people who have infection-derived immunity at the start of the next season. This is supported by statistical analyses indicating that susceptibility at the start of each season (based on the model fit in Dawa *et al*. (2020)) is not strongly dependent on infections in the previous season (Supplement section 4). To explore the possibility that there is some dependency, we run sensitivity analyses with two different assumptions on changes in susceptibility (Supplement section 9).

### Model 3 - Background FOI

To characterise influenza epidemiology in Kenya, we use weekly numbers of hospitalised patients with SARI from 2010-2018 from the Kenyan National SARI surveillance system (input 3). Data from a subset of 5 large hospitals that have a bed capacity of over 200 and a well-established surveillance system in place is used. The case definition of SARI was a hospitalised patient with acute illness onset presenting with fever or cough. A random sample of these patients underwent virological analysis to identify the presence or absence of influenza. For further details and data access, see Dawa *et al*. (2020).

To account for infections in the inter-epidemic periods, we include a background rate of infection with a Poisson distribution with shape parameter *Λ*_*i,k*_, fitted to the weekly observed cases in each age group and of each subtype across all inter-epidemic periods. We then calculate the weekly number of background infections per age group, *i*, and subtype, *k*, across the whole time period:

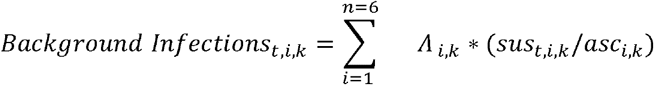

Where *sus*_*t,i,k*_ is the proportion susceptible each week (*t*) for age group *i* and influenza subtype *s* outputted from the vaccination model, *asc*_*i,k*_ is the mean ascertainment rate by age group as estimated in the Dawa *et al*. (2020) paper.

### Model 4 - Economic analyses

We estimate the incremental cost-effectiveness of each of the vaccine scenarios in Table 1 (compared to no vaccination), following WHO recommendations for economic evaluations of vaccines (14). The analytic time horizon used in the economic analyses is the same as the epidemiological model (2010-2019 inclusive), except that life years lost due to death are counted until the full normal life expectancy. Information on input costs used in these analyses (input 4) can be found in the Supplement section 8. We adopt a societal perspective on costs, and both costs and health outcomes are discounted at 3% per annum, with 0% discounting for health outcomes in a sensitivity analysis. All costs (except vaccine costs) are expressed in terms of 2019 USD and costs from other years are adjusted using Kenya gross domestic product (GDP) deflator values (15) before calculating cost-effectiveness measures.

Uncertainty is captured using probabilistic sensitivity analysis. This is done by drawing 1000 random samples per vaccine scenario of the total number of influenza infections generated from 2010 to 2019 by all virus subtypes across all age groups and charting disease and hospitalisation outcomes for each infection. Adopting the same approach as Dawa *et al*. (2020), we use a decision tree (Supplement section 8) to project health-related outcomes associated with influenza infections. Samples of probability parameters are drawn from a beta distribution (16) whose shape parameters were calculated first by fitting the mean and 95% confidence intervals for each probability parameter (drawn from the literature) to a beta distribution (2).

We further divide symptomatic infections into mild (upper respiratory tract infections, URTI) or severe (lower respiratory tract infections, LRTI) illness. Patients with mild illness will receive medical attention at outpatient clinics and eventually recover. Severely ill patients go on to be hospitalised, and further progress to recover from illness or die. The mean durations of influenza-associated illness and length of hospital stay are assumed to be 4 days (17,18).

A range of influenza-related healthcare utilisation events such as seeking medical care at outpatient clinics, hospitalisation as inpatients and purchase of over-the-counter medication are assumed to incur healthcare costs. To capture uncertainty around these costs, random samples of cost parameters are drawn from a gamma distribution (16). Direct medical costs include the price of influenza vaccines, assumed to be $3 per dose, and vaccine wastage, assumed to be 15% (2). Healthcare related costs include transportation costs for hospital visits to seek medical care for influenza-associated illness or for influenza vaccination. Similarly, indirect costs include lost wages and childcare costs due to influenza-related illness (see supplement section 8, and Figure 1 and Tables 2 and 3 in Dawa *et al*., (2020) for parameter values and references).

**Table 2:** Selected willingness to pay (WTP) thresholds used in this study.

| WTP threshold (USD) | Description* | Reference |
| --- | --- | --- |
| 100 | WHO best buy | (21) |
| 623 | 45% Kenya pc GDP (2015) | (19) |
| 1912.65 | 1x Kenya pc GDP (2019) | (22) |
| 5737.95 | 3x Kenya pc GDP (2019) | (22) |

\*pc GDP - per capita gross domestic product

**Table 3:** Median (and 95% CrI) values of threshold per-dose vaccine prices (2019 USD) at or below which each vaccination scenario is cost-effective, calculated using discounted costs and DALYs, at four selected thresholds of willingness-to-pay per DALY averted. These are calculated while including a median vaccine administration cost of $1.31 per dose (gamma distributed).

| **Vaccine** | **WHO best**<br>**buy (\$100)** | **45% per capita**<br>**GDP (\$623)** | **1x per capita**<br>**GDP (\$1913)** | **3x per capita**<br>**GDP (\$5738)** |
| --- | --- | --- | --- | --- |
| **Current**<br>**seasonal** | -0.87 (-3.74,<br>1.85) | -0.14 (-2.92,<br>2.63) | 1.54 (-1.43,<br>5.79) | 6.51 (1.54,<br>16.66) |
| **Improved**<br>**(minimal)** | -0.58 (-3.48,<br>3.29) | 0.45 (-2.49,<br>4.38) | 2.86 (-0.69,<br>8.77) | 9.97 (3.52, 23.2) |
| **Improved**<br>**(breadth)** | 0.98 (-2.05,<br>10.85) | 3.77 (0.03,<br>14.02) | 10.4 (4.05, 25.4) | 29.47 (13.83,<br>64.04) |
| **Improved**<br>**(efficacy)** | 0.46 (-2.42,<br>8.38) | 2.67 (-0.71,<br>10.9) | 7.99 (2.69,<br>19.89) | 23.31 (10.78,<br>50.57) |
| **Universal** | 1.59 (-1.51,<br>14.2) | 5.16 (0.94,<br>18.57) | 13.67 (5.99,<br>31.68) | 37.8 (18.74,<br>79.06) |

\*pc GDP - per capita gross domestic product

For health outcomes, we calculate disability-adjusted life years (DALYs) using disability weights for mild URTI, moderate and severe LRTI and death (GBD, 2019). In contrast to Dawa et al. (2020), no age-weighting of DALYs is done, as this is no longer recommended (14).

We determine cost-effectiveness of vaccination scenarios by calculating median incremental cost-effectiveness ratios (ICERs) per DALY averted and median incremental net monetary benefits (INMBs) across all ten years for each vaccine scenario compared to the no vaccination scenario. The most cost-effective scenario is the one with the lowest ICER value and the highest INMB value. In the absence of locally-determined cost-effectiveness thresholds for health interventions in Kenya, ICERs are evaluated against a WHO ‘best buy’ threshold of $100 per DALY averted in LMICs as well as cost-effectiveness thresholds derived using two broad approaches - marginal productivity thresholds calculated by the University of York (19) and those based on global analyses by the Commission for Macroeconomics and Health (20). While results using four WTP thresholds (Table 2) are presented in the main paper, details of the full range of thresholds used and corresponding results are presented in the supplement (section 8). These thresholds are also used to calculate vaccine prices at or below which a vaccination scenario is deemed cost-effective.

All results presented in the main text are calculated using discounted costs and DALYs. In sensitivity analyses, undiscounted costs are also used. We also analyse the effect of changing the vaccine price to $1.50, $6 and $10 per dose.

## Results

### Cases averted and doses used

While all modelled vaccine types increased the proportion of the population with some immunity, *Universal vaccines* resulted in the highest levels of immunity across the whole period (Supplemental section 7). In addition, this resulting immunity was generated with fewer vaccine doses due to slower waning, with a total of 14 million vaccine doses used for the *Universal vaccine* scenario over the whole time period. The same number of vaccines were used for *Improved vaccine (breadth*) scenarios, 19 million for *Improved vaccine (efficacy)* scenarios and 30 million for the *Current seasonal vaccines* and the *Improved vaccines (Minimal)* scenario.

The high immunity from *Universal* vaccines translated into the biggest projected reduction in cumulative infections across the 10 year period with a median total of 66% of infections averted (95% Credible Interval (CrI) 56%-74%) as compared to the no vaccination scenario. This compared to the *Improved (Efficacy)* of 57% (95% CrI 47 - 67%), *Improved (Breadth)* of 51% (95% CrI 42%-61%), *Improved (Minimal)* 41% (95% CrI 33% - 49%) and *Current seasonal* of 29% (95% CrI 23% - 35%) infections averted. The mean *R*_*0*_ of influenza across epidemics was 2.2 (range 1.2 - 6.7, Supplement section 8 for further details) and across vaccination scenarios the average number of cases averted per vaccine dose ranged from 0.33 to 2.6.

### Cost-effectiveness

Programmes using *Universal* and *Improved (Breadth) vaccines* incurred the lowest total vaccine purchase and administration costs across the entire period, assuming per-dose vaccine costs are the same for all vaccines ($3), because they required the fewest doses. These amounted to a median total value of $78.86 million (95% CrI: $60.96, $125.18 (in millions)), compared to $108.54 million for *Improved (Efficacy)* and $167.91 million for both *Improved (Minimal)* and *Current seasonal* (Supplement section 8). After accounting for these costs and the costs of travel to seek vaccination, programmes using universal vaccines incurred the lowest total societal costs (direct medical, healthcare-related and indirect costs) and thereby incremental total costs, compared to when no vaccination was conducted (Figure 3A). Median discounted incremental total costs for *Universal* vaccines were $27.67 million (95% CrI: $-174.38, $78.21 (in millions)). In contrast, median discounted incremental costs were higher for all *Improved* vaccines and highest for *Current seasonal* vaccines ($128.64 million (95% CrI $35.62, $228.43 (in millions)) (Supplement section 8).

**Figure 3:**
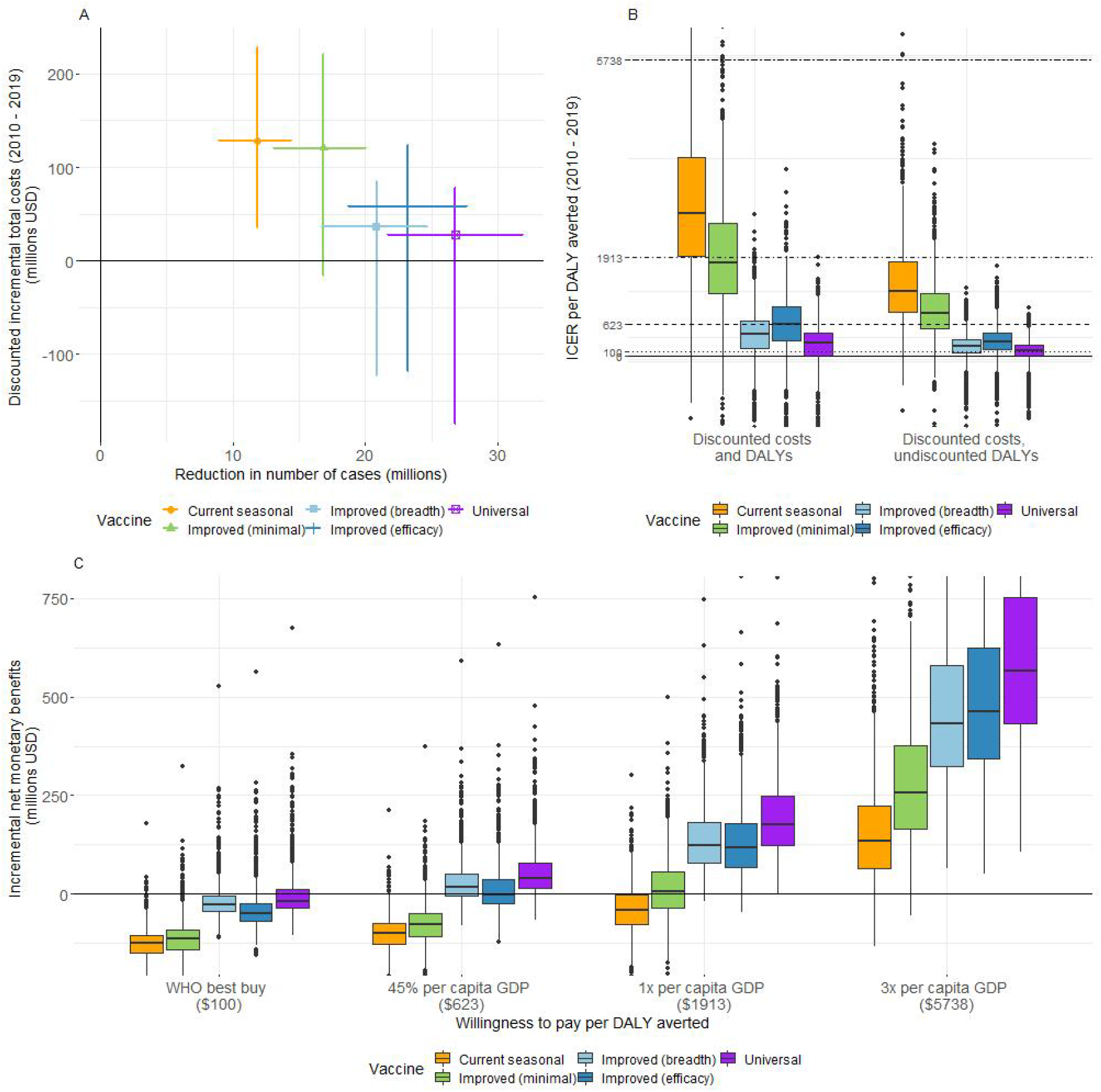
A) Mean (with 95% CrI) discounted incremental total costs (in millions of USD) vs. mean (with 95% CrI) reduction in number of cases (in millions) for each vaccine (2010 – 2019). B) Boxplot of ICER per DALY averted for each vaccine (2010 - 2019). Horizontal lines represent different willingness-to-pay thresholds per DALY averted. C) Boxplot of INMB (in millions of USD) (2010 to 2019) at four selected thresholds of WTP per DALY averted.

None of the vaccines were cost-effective at the WHO best buy threshold of $100 per DALY averted when evaluating cost-effectiveness using discounted costs and DALYs (Figure 3). While there was overlap between uncertainty ranges of ICER values calculated for all five vaccines, *Universal* vaccines were cost-effective across three of the four WTP thresholds evaluated in this study, with a median ICER per DALY averted of $263 (95% CrI: $-1698, $1061) (Figure 3B). Similarly, *Improved (breadth)* vaccines were cost-effective across three of four WTP thresholds with a median ICER value of $422 per DALY averted, while *Improved (Efficacy)* vaccines had a median ICER value of $626, being cost-effective across two of four thresholds. In contrast, *Current seasonal* vaccines had a median ICER value of $2764 per DALY averted, being cost-effective only at a WTP threshold of 3 times the 2019 per capita GDP of Kenya of approximately $5738 (Figure 3B, Supplement section 8). Thus, median ICER values for *Improved (Breadth), Improved (Efficacy)* and *Current seasonal* vaccines were 1.60, 2.38 and 10.51 times higher than for *Universal* vaccines, respectively.

Similarly, *Universal* vaccines had the highest median INMB values across all WTP thresholds (Figure 3C). At a threshold of $623 (45% of Kenya’s 2019 per capita GDP), the median INMB value of *Universal* vaccines ($39.6 million) was 2.29 times higher than that of *Improved (Breadth)* ($17.26 million) vaccines (Supplement section 8). At this threshold, *Universal* vaccines had a high probability (>75%) of being cost-effective, at or below a median price of $5.16 per vaccine dose (95% CrI: $0.94, $18.57) (Table 3, Fig. S7, supplement section 8). Calculated threshold per-dose vaccine prices were consistently higher for *Universal* vaccines across all WTP thresholds. *Universal* vaccines had median INMB values 4.21 times higher than that of *Current seasonal* vaccines ($134.63 million) at a WTP threshold of $5738.

In our sensitivity analyses, increased coverage of vaccination made only slight differences to the cost-effectiveness of any of the vaccines across the different WTP thresholds evaluated (supplement, section 9). The number of cases averted per vaccine dose was slightly lower than in the 50% coverage scenario, ranging from 0.31 - 2.16. In addition, we found that assumptions around susceptibility had a large impact on impact and cost-effectiveness. If we assumed that greater reduction in infections in one season increased susceptibility in the next season, then vaccines were less impactful and cost-effective (see supplement section 10 for details).

## Discussion

Our study indicates that next generation vaccines are likely to have much greater impact and an improved cost-effectiveness profile than currently available influenza vaccines. This is true even for incrementally improved vaccines with slightly greater breadth or duration. These are evidenced by the scale of reduction in influenza infections and improvements in cost-effectiveness measures, particularly for universal vaccines. Universal vaccines result in the most substantial reduction in influenza infections utilising the least vaccine doses, averting 66% of infections compared to no vaccination. In contrast, our model predicts that current vaccines avert only 29% of infections, while even improved (minimal) vaccines avert 41% of infections. Similarly, ICER values are higher for improved (1.60 to 2.38 times) and current seasonal (10.51 times) vaccines than for universal vaccines. Universal vaccines also have the highest INMB values - 2.29 times higher than Improved (breadth) vaccines at a WTP threshold of $623, and 4.21 times higher than current seasonal vaccines at a WTP threshold of $5738, the only threshold at which current vaccines are cost-effective. Thus our results suggest that universal vaccines result in the highest immunity per vaccine dose and subsequently, the least number of infections, as well as having the most favourable cost-effectiveness profile among all the vaccines evaluated.

Our conclusions are influenced by vaccine dose costs and cost-effectiveness thresholds. We assumed that vaccine per-dose costs are the same for all vaccines ($3), which is unlikely to be true. More advanced vaccines may cost more per dose. As a comparison, SARS-Cov-2 AstraZeneca vaccines, which were sold without profit, cost between $2.15 and $5.25 per dose, compared to mRNA SARS-CoV-2 vaccines costing $14.70 to $23.50 per dose (23). Nevertheless, our estimates of the threshold per-dose vaccine price (prices at or below which vaccination programs are cost-effective) suggest that universal vaccines are cost-effective even when priced higher than current seasonal or improved vaccines and irrespective of the WTP threshold. At the same time, we find that improved vaccines can also be cost-effective at comparatively low WTP thresholds and result in fewer influenza cases than currently available seasonal vaccines, even if priced higher per dose. The development and use of universal vaccines are very likely to benefit low-and-middle-income countries which may only be willing or able to pay less for health benefits than more advanced economies. However, universal vaccines are unlikely to be immediately available for widespread use, but improved vaccines offer substantial value as an achievable and satisfactory alternative to current influenza vaccines, especially since these may be available in the near future.

Our analyses demonstrate the importance of assumed cost-effectiveness thresholds when determining whether health interventions are cost-effective or not. Kenya does not have an official cost-effectiveness threshold, but Dawa *et al*. (2020) reported that vaccination with current seasonal influenza vaccines in Kenya had a low probability of being cost-effective given WTP thresholds of 1-51% of per capita GDP. To address uncertainty around thresholds, we used a wide range of values ranging from extremely low WHO “best buys” threshold reserved for evaluating some of the most cost-effective programmes that WHO has ever evaluated (21), to very high 1-3 times GDP per capita thresholds representing the potential value of human capital associated with disability (24). Like Dawa *et al*., we find that current vaccines are cost-effective only at a very high threshold of 3 times the per capita GDP of Kenya and at a maximum threshold price of $6.51 per dose, which is much lower than prices at which most influenza vaccines are available in the US (25) or UK (26). Conversely, both universal and improved influenza vaccines are cost-effective at lower thresholds.

A key strength of our epidemiological model is the direct incorporation of vaccine-derived immunity waning over multiple years, with ageing of the population, which is required to evaluate next generation vaccines with benefits that last several years. This contrasts to many seasonal vaccination models where vaccine-derived immunity is not tracked across seasons (2,13). In contrast with the marked annual seasonality of influenza in temperate regions (27), influenza epidemics in Kenya do not have a regular seasonal pattern, with substantial transmission in between epidemics, which we included by separately modelling the inter-epidemic periods. However we used a relatively simple approach for this and we do not capture indirect effects of vaccination between epidemics. In addition, while we include 3 influenza subtypes (AH1N1, A H3N2 and B), we do not allow for any interaction between these subtypes, which may contribute to the dynamics of transmission (28–33). However, as our modelling is based on fitted models, this should not have major impacts on our economic analysis. Therefore the main practical disadvantage is that we are unable to investigate vaccines with different efficacies within the influenza B viruses.

Country decisions to invest in health interventions can be influenced by considerations other than cost-effectiveness (34), for example due to competing options for implementation or due to widespread vaccine hesitancy. Whilst our modelling indicates that next generation vaccines can be cost-effective, their implementation will be competing against other public health interventions. In Kenya, separate studies have shown both rotavirus and pneumococcal childhood vaccination to be cost effective, with between $25 and $59 (35) and $38 (36) per DALY averted respectively, and programmes covering these vaccines have been introduced. However, these estimates are substantially lower than for even the universal influenza vaccines calculated here. Equity in vaccine distribution (37) is also a key consideration for vaccine programme implementation. The availability of financing options such as from Gavi, the Vaccine Alliance (38), is also important, but Kenya is already starting to transition out of Gavi support.

Our study has a number of other limitations. We have assumed that vaccination occurs independent of current vaccine status, meaning that individuals can receive multiple vaccinations and therefore some vaccinations will be ‘wasted’ on individuals already immune. This is a conservative assumption, likely making the vaccine scenarios appear less cost-effective, and is more likely to have an effect at higher coverage levels. It is also recommended that children between 6 months and 8 years of age, or those who have only ever received one dose, should receive two vaccine doses at least 4 weeks apart (39,40). Administration of a second vaccine dose will incur additional costs for vaccine purchase, transport and administration, although these additional costs may be off-set by vaccinating independent of vaccine status. In reality, there may also be challenges to administer vaccines twice due to limited access. We also do not consider adverse vaccine reactions (40,41) in our DALY calculations. These would influence cost-effectiveness and vaccine threshold prices, particularly at lower cost-effectiveness thresholds.

Immune protection to influenza virus infection and vaccination are poorly understood and we found that assumptions on infection-derived immunity have a large impact on incidence and resulting cost-effectiveness estimates. However, such assumptions could not be empirically informed, because in this setting the previous season does not have an impact on estimated susceptibility levels in the following season and our sensitivity analyses with different infection-susceptibility assumptions show different behaviour than observed for current seasonal vaccines. Therefore our main analysis presents the most likely assumptions. Another important consideration is the potential population-level effects of universal vaccines on vulnerability to newer influenza virus variants. Previous mathematical modelling studies suggest that universal vaccines can prevent the development of cross-protective immunity developed through natural infection. In the absence of sufficiently high vaccination coverage, it was thus suggested that universal vaccines can increase the risks of emergence of vaccine escape variants that could cause influenza pandemics (42,43). These studies suggest that combining administration of seasonal and universal vaccines may help to mitigate these risks (42), a strategy which we have not explored in our study.

## Conclusions

Our study provides the first formal evaluation incorporating both direct and indirect (herd) protection, of the effectiveness and cost-effectiveness of a range of next generation influenza vaccines meeting WHO PPCs. In doing so it bolsters the case for investing in development of these vaccines, while highlighting the benefits to be derived from improved vaccines. This provides proof-of-principle for similar studies to be conducted in other LMICs, so that a global picture of potential demand for these vaccines can be built.

## Data Availability

All data used is available in the Supplement of Dawa et al. (2020).

https://bmcmedicine.biomedcentral.com/articles/10.1186/s12916-020-01687-7#Sec18

## List of Abbreviations

CrI: Credible Interval
DALY: disability-adjusted life years
E: Exposed
GDP: gross domestic product
HA: haemagglutinin
I: infectious
ICER: incremental cost-effectiveness ratio
INMB: incremental net monetary benefits
LMIC: lower middle-income countries
LRTI: lower respiratory tract infections
NH: Northern Hemisphere
NIAID: National Institute of Allergy and Infectious Diseases
PPC: Preferred product characteristics
R: recovered
Rv: Recovered-vaccinated
S: Susceptible
SARI: severe acute respiratory illness
SH: Southern Hemisphere
Sv: Susceptible-vaccinated
URTI: upper respiratory tract infections
USD: US Dollars
VE: Vaccine efficacy
WHO: World Health Organisation
WTP: Willingness-to-pay

## Declarations

### Ethics Approval

Not applicable.

### Consent for publication

Not applicable

### Availability of data and materials

The datasets analysed during the current study are available in the Supplement of Dawa *et al*. (2020). (https://bmcmedicine.biomedcentral.com/articles/10.1186/s12916-020-01687-7#Sec18)

All code is available at https://github.com/NaomiWaterlow/NextGenFlu_Kenya.

### Authors contributions

RME, MJ, JB and MM contributed to the conception of the project. RME, MJ, JD, NRW, SR and EvL contributed to the design of the work. NRW, SR, JD and EvL contributed to the analysis of the work. NRW, SR, JD, EvL, JB, MM, PL, RME and MJ contributed to the interpretation of the data. NRW, SR and JD contributed to the creation of the new software in the work. NRW and SR drafted the manuscript and NRW, SR, JD, EvL, JB, MM, PL, RME and MJ reviewed and edited the manuscript.

## Acknowledgments

We would like to acknowledge the advice received from Sandra Chaves, Marc-Alain Widdowson and Gideon Emukule.

