## Supplemental File for "Potential health and economic impact of paediatric vaccination using next generation influenza vaccines in Kenya: a modelling study"

Contents:

1. Vaccination model equations
2. Epidemic model equations
3. Vaccine efficacy and vaccine match/mismatch by season
4. Epidemic timings
5. Relationship between infection in previous season and susceptibility in current season
6. Cases and Immunity by age

### 7. Basic reproduction number for each epidemic in 2010-2018

8. Further details of cost-effectiveness analyses
9. Sensitivity analyses
10. References

### 1. Vaccination model equations

The vaccination model equations (Model 1 in Figure S1) are shown below. This model is used to track immunity over time. The epidemics are modelled separately (Section 2 below, Model 2 in Figure S1). This model has three compartments, Susceptible and unvaccinated (S), Susceptible and Vaccinated (Sv) and Immune and vaccinated (Rv). The proportion vaccinated that move into the immune vs susceptible compartments are determined by the vaccine efficacy ($\alpha$).
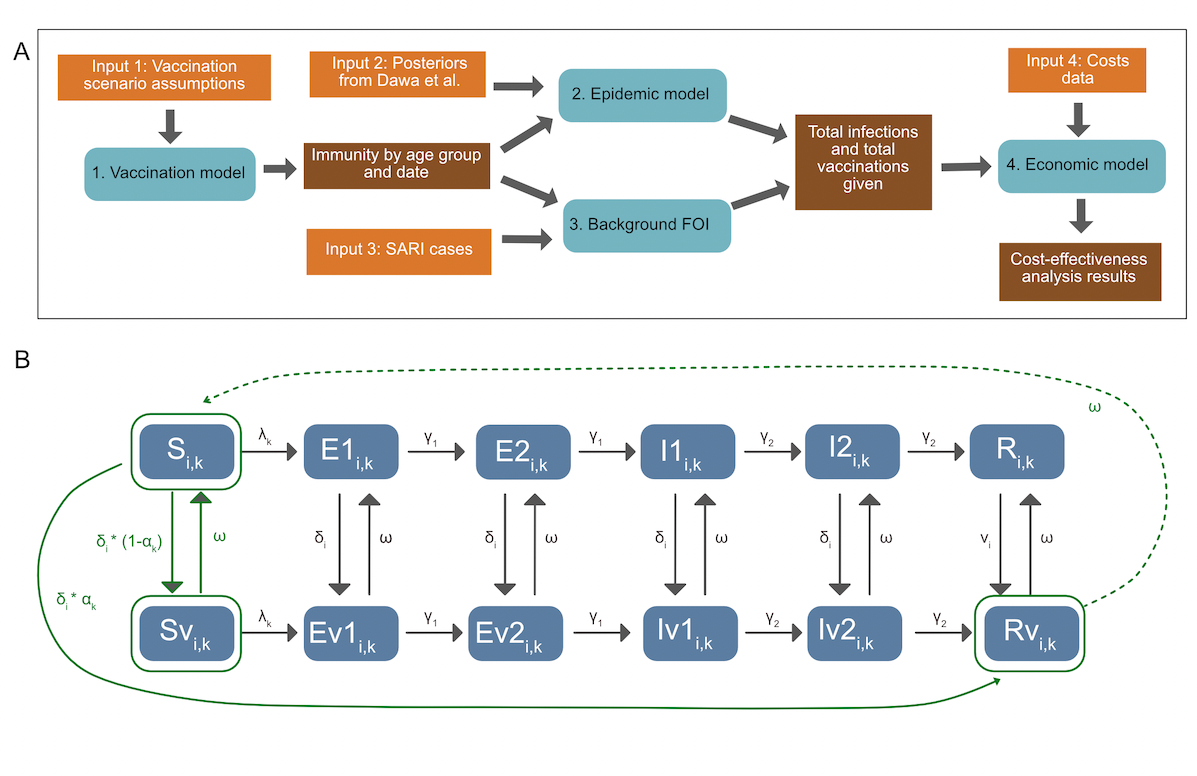

Figure S1: *Modelling overview: A) Methods overview, depicting inputs, models and outputs. B) Model diagram, including both the epidemic and the vaccination model. Elements in solid green are included in both models. Transitions in grey are included only in the epidemic model, and transitions in dotted green are included only in the vaccination model. States are: Susceptible (S), Exposed (E), Infectious (I) and Recovered (R), and their vaccinated counterparts (Sv, Ev1, Ev2, Iv1, Iv2, Rv). v denotes the vaccinated equivalent of the compartments. See Table S3 for parameter details.* $\delta$ *is the rate of vaccination in age group* $i$*,* $\alpha$ *is the efficacy by subtype (*$k$*),* $\omega$ *is vaccine derived immunity waning. The model is run separately for each subtype. For the epidemic model, in both vaccinated and unvaccinated compartments, susceptibles who are infected with the viral subtype enter the first Exposed (E) compartment. They then progress through the E and Infectious (I) compartments. After ceasing to be infectious they enter the R compartment, whereupon they cannot be re-infected during the same epidemic period. Both the E and I populations consist of two compartments, in order to get a gamma distributed waiting time. Each compartment is also subdivided by age (i).*

**Vaccination Model Equations (Model 1)**

$$\frac{{dS}_{i}}{dt} = - \delta_{i}*S_{i} + \omega*({Sv}_{i} +{Rv}_{i})$$

$$\frac{{dSv}_{i}}{dt} = \delta_{i}*S_{i} * {(1-\alpha}_{i})- \omega*{Sv}_{i}$$

$$\frac{{dRv}_{i}}{dt} = \delta_{i}*S_{i} * \alpha_{i}- \omega*{Rv}_{i}$$

Subscripts:

$i$ - age group

States:

$S$ - Susceptible

$Sv$ - Vaccinated and susceptible

$Rv$ - Vaccinated and immune

Parameters:

$v$ - vaccination rate

$\alpha$ - vaccine efficacy

$1/\omega$ - duration of vaccine immunity

In addition vaccines used are tracked by the following equation where ${pop}_{i}$ represents the age-group specific population sizes.

$$\frac{{dV}_{i}}{dt} = v_{i}*{pop}_{i}$$

As in Dawa et al we estimate the annual population in each age group assuming a constant growth rate between 2010 and 2015 and then apply the World Bank annual population growth rates for subsequent years(1).

### 2. Epidemic model equations

The epidemic model equations are shown below, corresponding to Model 2 in Figure S1. From both vaccinated and unvaccinated compartments, susceptibles who are infected with the viral subtype enter the first Exposed (E) compartment. They then progress through the E and Infectious (I) compartments. After ceasing to be infectious they enter the R compartment, whereupon they cannot be re-infected during the same epidemic period. Both the E and I populations consist of two compartments, in order to get a gamma distributed waiting time. Each compartment is also subdivided by age (i). The model diagram is shown in Figure S1B. The model is run separately for each season and subtype. Parameter definitions are shown in table S1.

Force of Infection:

$\lambda_{i} = \beta*\zeta_{i}*\sum_{j=1}^{j=N} c_{ij}*({I1}_{j}+{I2}_{j}+{Iv1}_{j}+{Iv2}_{j})$

Ordinary differential equation model:

$$\frac{{dS}_{i}}{dt} = - \lambda_{i}*S_{i}- v_{i}*S_{i} + \omega*({Sv}_{i} +{Rv}_{i})$$

$$\frac{{dE1}_{i}}{dt} = \lambda_{i}*S_{i}-\gamma_{1}*{E1}_{i}- \delta_{i}*{E1}_{i} + \omega*{Ev1}_{i}$$

$$\frac{{dE2}_{i}}{dt} = \gamma_{1}*{E1}_{i}-\gamma_{1}*{E2}_{i}- \delta_{i}*{E2}_{i} + \omega*{Ev2}_{i}$$

$$\frac{{dI1}_{i}}{dt} = \gamma_{1}*{E2}_{i}-\gamma_{2}*{I1}_{i}- \delta_{i}*I1 + \omega*{Iv1}_{i}$$

$$\frac{{dI2}_{i}}{dt} = \gamma_{2}*{I1}_{i}-\gamma_{2}*{I2}_{i}- \delta_{i}*I2 + \omega*{Iv2}_{i}$$

$$\frac{{dR}_{i}}{dt} = \gamma_{2}*{I2}_{i}- \delta_{i}*R_{i} + \omega*{Rv2}_{i}$$

$$\frac{{dSv}_{i}}{dt} = - \lambda_{i}*{Sv}_{i}+ (1-\alpha_{i})*\delta_{i}*S_{i} - \omega*{Rv}_{i}$$

$$\frac{{dEv1}_{i}}{dt} = \lambda_{i}*{Sv}_{i}-\gamma_{1}*{Ev1}_{i}+ \delta_{i}*{E1}_{i} - \omega*{Ev1}_{i}$$

$$\frac{{dEv2}_{i}}{dt} = \gamma_{1}*{Ev1}_{i}-\gamma_{1}*{Ev2}_{i}+ \delta_{i}*{E2}_{i} - \omega*{Ev2}_{i}$$

$$\frac{{dIv1}_{i}}{dt} = \gamma_{1}*{Ev2}_{i}-\gamma_{2}*{Iv1}_{i}+\delta_{i}*I1 - \omega*{Iv1}_{i}$$

$$\frac{{dIv2}_{i}}{dt} = \gamma_{2}*{Iv1}_{i}-\gamma_{2}*{Iv2}_{i}+\delta_{i}*I2 - \omega*{Iv2}_{i}$$

$$\frac{{dRv}_{i}}{dt} = \gamma_{2}*{Iv2}_{i}+\delta_{i}*R_{i}+{\alpha_{i}*\delta_{i}}*S_{i}- \omega*{Rv2}_{i}$$

*Table S1: Parameters used in epidemic models*

| **Parameter** | **Symbol** | **Type (Fixed posterior vaccine assumption)** | **Value (if fixed)** |
| --- | --- | --- | --- |
| Age-specific vaccination rate | $\delta_{i}$ | Vaccine assumption | - |
| Vaccine efficacy | $\alpha$ | Vaccine assumption | - |
| Age specific force of infection | $\lambda_{i}$ | Posterior | - |
| Transmission rate | $\beta$ | Posterior | - |
| Contact rates between age groups i and j | $c_{ij}$ | Fixed based on Kiti *et al.* (2014)(2) | See paper |
| Latency period | $2 *1/\gamma_{1}$ | Fixed *fluEvidenceSynthesis* package | 0.8 days |
| Infectious Period | $2 *1/\gamma_{2}$ | Fixed *fluEvidenceSynthesis* package | 1.8 days |
| Vaccine immunity duration | $\omega$ | Vaccine assumption | - |
| Age specific proportion in vaccinated compartments at start of epidemic | ${\eta v}_{i}$ | Vaccine assumption / Model | - |
| Age specific proportion in Rv vs Sv compartments at start of epidemic | ${\eta Rv}_{i}$ | Vaccine assumption / Model | - |
| Age-specific susceptibility (<15 15-49 50+) | $\zeta_{i}$ | Posterior adapted to take into account previous seasons vaccination. | - |
| Age-specific ascertainment rates (<1 1-5 5+) | $\epsilon_{i}$ | Posterior | - |

### 3. Vaccine efficacy and vaccine match/mismatch by season

Table S1 shows the vaccine efficacy by year, hemisphere and viral subtype, and whether it is considered a match. If vaccine efficacy >= 50% the vaccine is considered a match to circulating strain and if vaccine efficacy < 50% the vaccine is considered a mis-match to the circulating strain as in Dawa et al. (2010).

*Table S2: Vaccine efficacy and vaccine match/mismatch by season. Acronyms are: Vaccine Efficacy (VE) Reference (Ref) NH (Northern Hemisphere) Southern Hemisphere (SH). * values shown represent VE against all subtypes. ** no VE values available for this period as such VE values from the preceding NH vaccine are used. *** measured only in children. **** very low cases assuming the VE is the same as the other strain. If VE >= 50% the vaccine is considered a match to circulating strain (Y) if VE < 50% the vaccine is considered a mis-match to the circulating strain (N) as in Dawa et al. (2010).*

| **Year** | **Hemisphere** | **AH1N1** | | | **AH3N2** | | | **B** | | |
| --- | --- | --- | --- | --- | --- | --- | --- | --- | --- | --- |
|  |  | **VE** | **Match?** | **Ref** | **VE** | **Match?** | **Ref** | **VE** | **Match?** | **Ref** |
| **2009** | **NH** | 62% | Y | (3) | 62%**** | Y | (3) | 62%**** | Y | (3) |
| **2010** | **SH** | 72%* | Y | (1) | 72%* | Y | (1) | 72%* | Y | (1) |
| **2010** | **NH** | 67% | Y | (4) | 67%**** | Y | (4) | 50% | Y | (1) |
| **2011** | **SH** | 72%* | Y | (1) | 72%* | Y | (1) | 72%* | Y | (1) |
| **2011** | **NH** | 65% | Y | (5) | 39% | N | (1) | 58% | Y | (5) |
| **2012** | **SH** | 65%** | Y | (5) | 39%** | N | (1) | 58%** | Y | (5) |
| **2012** | **NH** | 49%* | N | (3) | 47% | N | (3) | 67% | Y | (3) |
| **2013** | **SH** | 52%* | Y | (6) | 52%* | Y | (6) | 54% | Y | (1) |
| **2013** | **NH** | 54% | Y | (1) | 52%* | Y | (3) | 52% | N | (3) |
| **2014** | **SH** | 64%*** | Y | (7) | 6%*** | Y | (7) | 23%*** | N | (7) |
| **2014** | **NH** | 23%* | N | (3) | 13% | N | (3) | 23%* | N | (3) |
| **2015** | **SH** | 24% (all A) | N | (8) | 22% | N | (8) | 46% | N | (8) |
| **2015** | **NH** | 41% | N | (3) | 48%* | N | (3) | 55% | Y | (1) |
| **2016** | **SH** | 40%* | N | (9) | 40%* | N | (9) | 40* | N | (9) |
| **2016** | **NH** | 40%* | N | (3) | 43% | N | (3) | 73% | Y | (3) |
| **2017** | **SH** | 50% | Y | (10) | 10% | N | (10) | 57% | Y | (10) |
| **2017** | **NH** | 65% | Y | (3) | 25% | N | (3) | 42% | N | (1) |
| **2018** | **SH** | 65%** | Y | (3) | 25%** | N | (3) | 42%*8 | N | (1) |
| **2018** | **NH** | 67% | Y | (1) | 9% | N | (3) | 34% | N | (3) |

### 4. Epidemic timings

Table S3 defines the timings of the epidemic and inter-epidemic periods of the model, as well as the subtype and the year from which the population size is used. A total of 11 epidemics are included.

*Table S3: Epidemic and Inter-epidemic time periods.*

| **Start** | **End** | **Type** | **Subtype** | **Population** |
| --- | --- | --- | --- | --- |
| 2012-01-01 | 2013-12-20 | Inter-Epidemic | AH1N1 | - |
| 2013-12-20 | 2014-09-05 | Epidemic | AH1N1 | 2014 |
| 2014-09-05 | 2018-01-19 | Inter-Epidemic | AH1N1 | - |
| 2018-01-19 | 2018-10-12 | Epidemic | AH1N1 | 2018 |
| 2010-01-01 | 2010-03-12 | Inter-Epidemic | AH3N2 | - |
| 2010-03-12 | 2010-12-17 | Epidemic | AH3N2 | 2010 |
| 2010-12-17 | 2011-12-23 | Inter-Epidemic | AH3N2 | - |
| 2011-12-23 | 2012-05-11 | Epidemic | AH3N2 | 2012 |
| 2012-05-11 | 2012-05-11 | Inter-Epidemic | AH3N2 | - |
| 2016-03-25 | 2016-11-25 | Epidemic | AH3N2 | 2016 |
| 2016-11-25 | 2018-06-15 | Inter-Epidemic | AH3N2 | - |
| 2018-06-15 | 2018-12-14 | Epidemic | AH3N2 | 2018 |
| 2018-12-14 | 2010-12-17 | Inter-Epidemic | B |  |
| 2010-12-17 | 2011-08-05 | Epidemic | B | 2011 |
| 2011-08-05 | 2011-08-12 | Inter-Epidemic | B |  |
| 2011-08-12 | 2012-03-19 | Epidemic | B | 2011 |
| 2012-03-19 | 2013-05-03 | Inter-Epidemic | B |  |
| 2013-05-03 | 2013-12-13 | Epidemic | B | 2013 |
| 2013-12-13 | 2015-11-27 | Inter-Epidemic | B |  |
| 2015-11-27 | 2016-05-20 | Epidemic | B | 2016 |
| 2016-05-20 | 2017-09-01 | Inter-Epidemic | B |  |
| 2017-09-01 | 2018-06-22 | Epidemic | B | 2018 |

### 5. Relationship between infection in previous season and susceptibility in current season

We posit that vaccination may reduce the opportunity to acquire infection-derived immunity and hence increase susceptibility in individuals whose vaccine protection has waned compared to non-vaccinated people. In order to investigate this potential relationship we investigated the relationship between the number of infections in the previous season compared to the susceptibility in the current season. If there is a strong correlation between them it implies that infections in the previous season have a large impact on the current season and hence the susceptibility is strongly influenced by the infection-derived immunity in the previous season. Alternatively if there is no / a weak correlation then we can conclude that susceptibility is not strongly driven by infections in the previous season. We analysed this for both the previous subtype specific epidemic as well as for the previous epidemic regardless of subtype. We found that there was a very weak correlation for both scenarios with an R^2^ value of 0.034 and 0.023 respectively. Figure S2 shows the scatter plots broken down by age group.

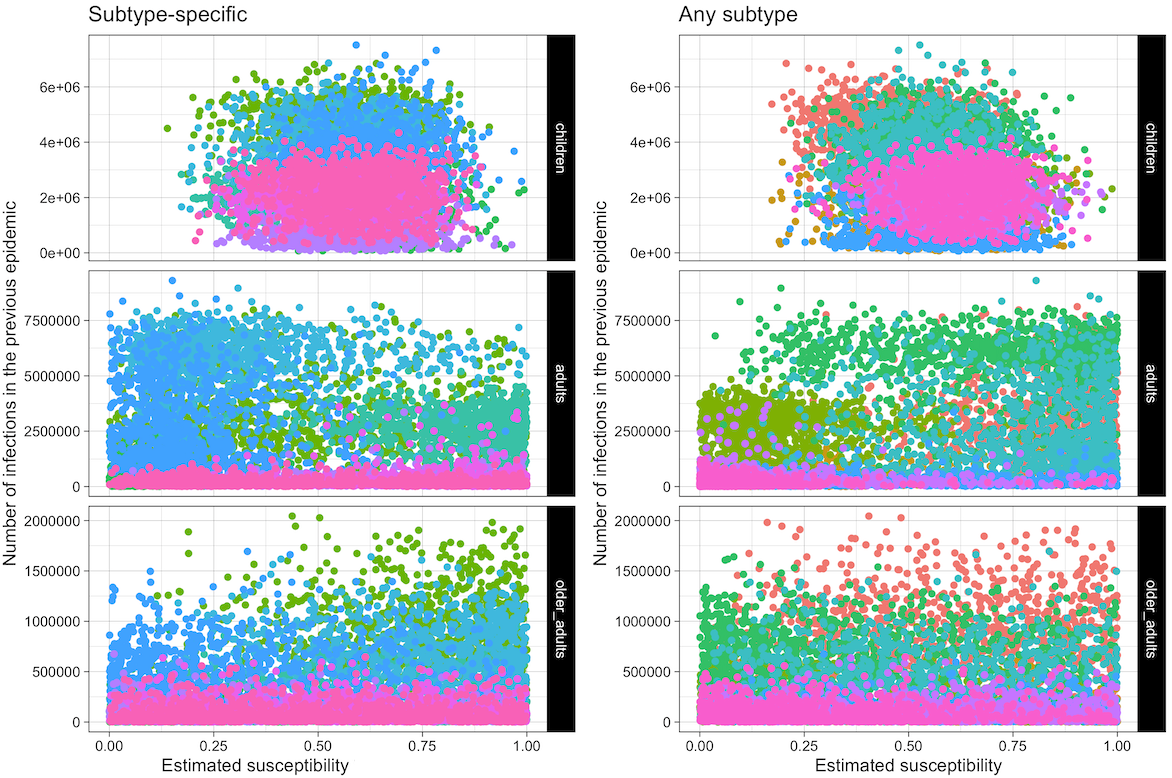

*Figure S2: Scatter plots showing the number of infections in the previous epidemic (either subtype specific or not) against the susceptibility values in the current epidemic. Each point represents one sample from the posterior and colours indicate epidemics.*

### 6. Cases and Immunity output by age and viral subtype

Figure S3 shows the vaccine-derived population immunity by age group and virus subtype for the model. Figure S4 shows the cases over time by scenario broken down by age group and viral subtype.

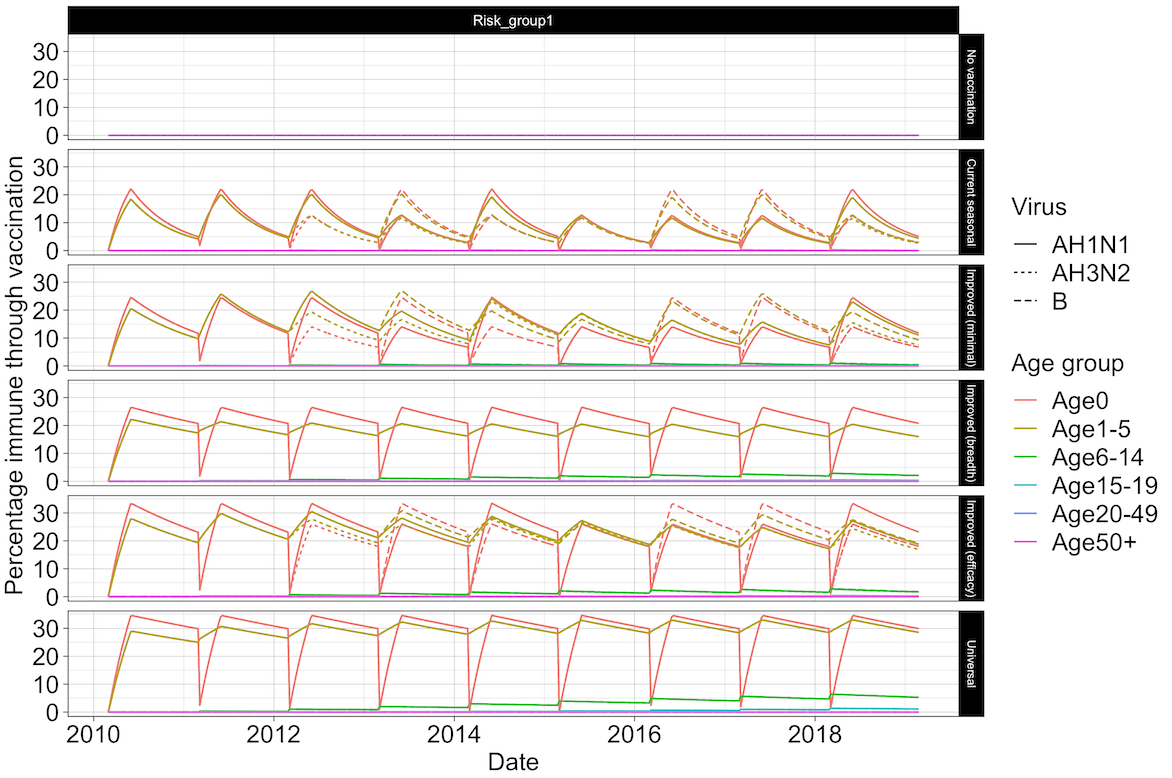

*Figure S3: Percentage of the population with vaccine immunity for each scenario split by virus subtype and age group.*

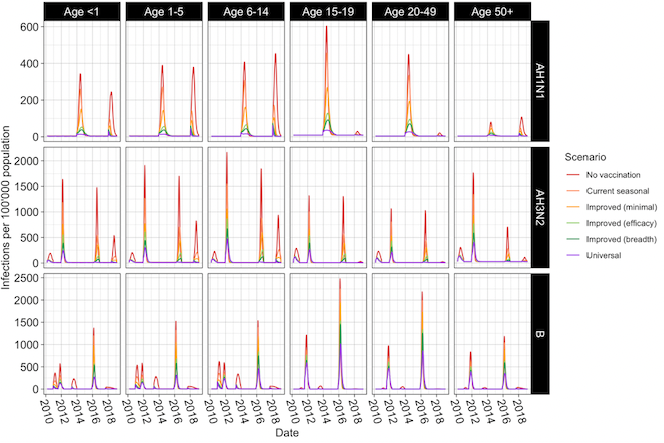
*Figure S4: Infections per 100’000 population by Age group and virus subtype, for each vaccine scenario.*

### 7. Basic reproduction number for each epidemic in 2010-2018

The basic reproduction numbers of each epidemic in the dataset are shown in Figure S5. These were calculated using the Next Generation Matrix method as in (11). Transmission ($T$) and Transition ($\Sigma$) matrices for just one age group are shown below, see github repository for the full matrices. $\beta$ is the transmission rate, $\alpha_{i,j}$ is the contact rate between age groups i and j, ${2*1/\gamma}_{1}$ is the latency period and ${2*1/\gamma}_{2}$ is the infectious period.

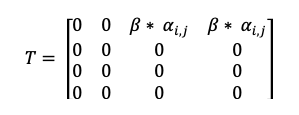

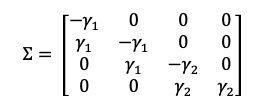

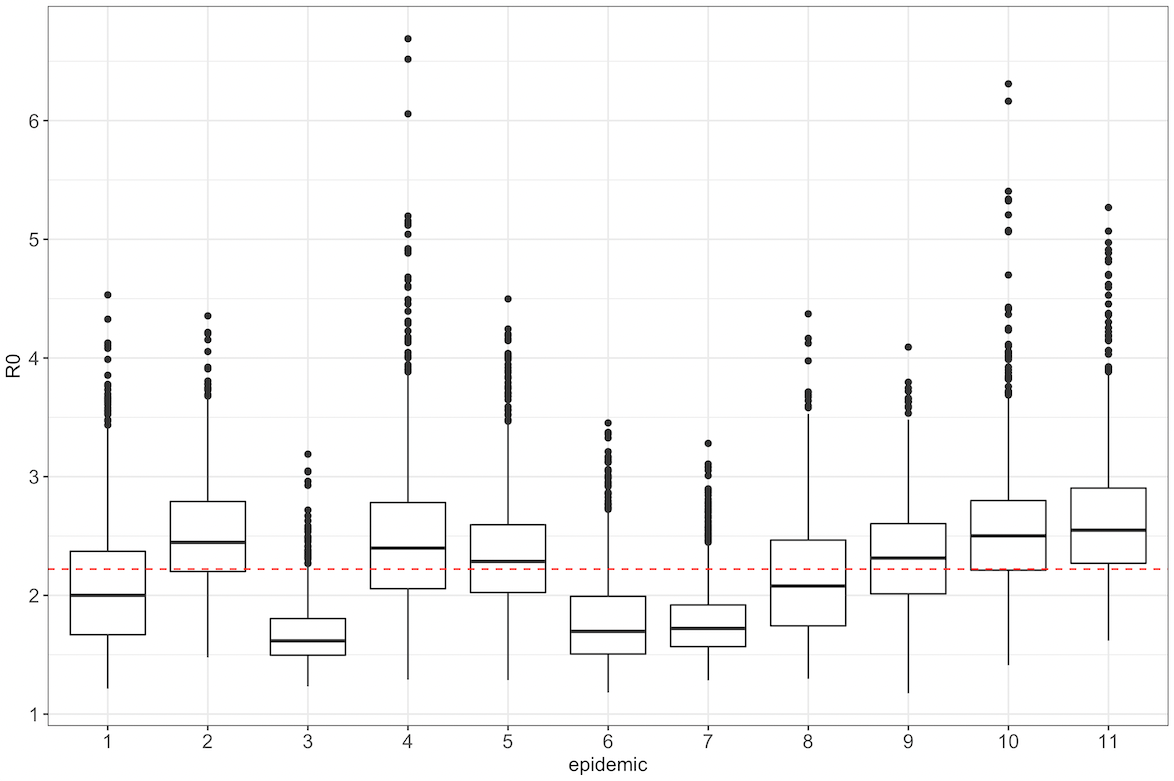

*Figure S5: Distribution of R_0_s across epidemics. The red dashed line indicates the means across all epidemics.*

### 8. Further details of cost-effectiveness analyses

8.1 Decision tree model

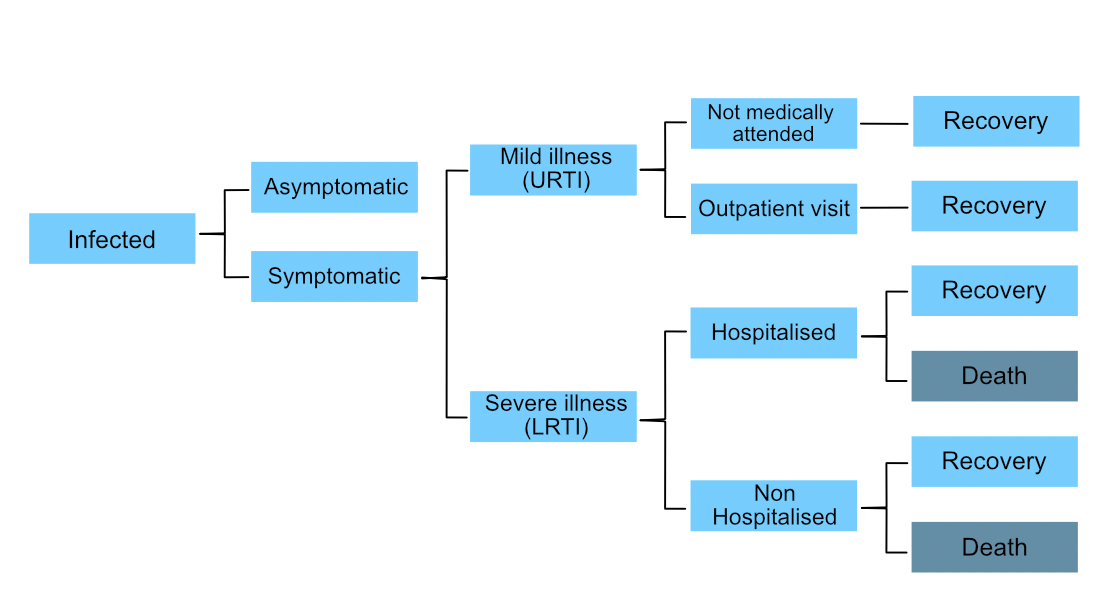

*Figure S6: Economic evaluation decision tree used to project health-related outcomes associated with influenza infections (adapted from Dawa et al. 2020* *(1)**). URTI stands for Upper Respiratory Tract Infection and LRTI for Lower Respiratory Tract Infection.*

*8.2 Input parameters*

*Table S4: Values for parameters governing disease state transitions healthcare utilisation and DALY weights used in the economic model. Means and 95% confidence intervals (CI) (or means only where confidence intervals are not available) are presented. Samples of these parameters were drawn from a beta distribution whose shape parameters were calculated first by fitting the mean and confidence intervals to a beta distribution*

| Outcome | Age group | Mean | Lower 95% CI | Upper 95% CI | Reference |
| --- | --- | --- | --- | --- | --- |
| **Parameters governing disease state transitions** | | | | | |
| Proportion developing any clinical illness | All ages | 0.669 | 0.583 | 0.745 | (12) |
| Proportion developing URTI | All ages | 0.588 | 0.455 | 0.708 |  |
| Proportion developing lower respiratory tract symptoms | All ages | 0.21 | 0.14 | 0.303 |  |
| Proportion of influenza hospitalised cases that die | <1 yr | 0.0274 | 0 | 0.0616 | (1) |
|  | 1-5 yrs | 0.0091 | 0 | 0.0322 |  |
|  | 6-14 yrs | 0.0108 | 0 | 0.0902 |  |
|  | 15-19 yrs | 0 | 0 | 0.1116 |  |
|  | 20-49 yrs | 0.0331 | 0 | 0.0818 |  |
|  | >50 yrs | 0.1818 | 0.0909 | 0.308 |  |
|  | All ages | 0.02 | 0.0035 | 0.0373 |  |
| Proportion of resp deaths that occur in hospitals | <1 yr | 0.2794 | 0.2451 | 0.314 | (1) |
|  | 1-5 yrs | 0.2899 | 0.2471 | 0.3349 |  |
|  | 6-14 yrs | 0.4361 | 0.3534 | 0.5278 |  |
|  | 15-19 yrs | 0.525 | 0.375 | 0.6795 |  |
|  | 20-49 yrs | 0.5067 | 0.4626 | 0.5525 |  |
|  | >50 yrs | 0.2715 | 0.2421 | 0.3012 |  |
|  | All ages | 0.3287 | 0.3106 | 0.3474 |  |
| **Parameters governing health care utilisation events** | | | | | |
| Proportion of outpatient cases who purchased medication prior to clinic visit | All ages | 0.718 | NA | NA | (13) |
| Proportion of hospitalised cases who sought care after discharge from hospital | All ages | 0.105 | NA | NA |  |
| Proportion of inpatient cases where household members missed work due to illness | All ages | 0.848 | NA | NA |  |
| Proportion of outpatient cases where household members missed work due to illness | All ages | 0.518 | NA | NA |  |
| Proportion of inpatient cases where household members paid for child care in the course of sickness | All ages | 0.029 | NA | NA |  |
| Proportion of outpatient cases where household members paid for child care in the course of sickness | All ages | 0.018 | NA | NA |  |
| Proportion of cases with influenza clinical illness who are hospitalised | 0-5 yrs | 0.0102 | 0.0089 | 0.0117 | (14) |
|  | 6-12 yrs | 0.0007 | 0.0006 | 0.001 |  |
|  | 13-17 yrs | 0.0006 | 0.0004 | 0.0011 |  |
|  | 18-24 yrs | 0.0008 | 0.0006 | 0.001 |  |
|  | 25-44 yrs | 0.0021 | 0.0018 | 0.0024 |  |
|  | 45-64 yrs | 0.0026 | 0.002 | 0.0033 |  |
|  | 65+ yrs | 0.0033 | 0.0025 | 0.0044 |  |
| Proportion of cases with influenza clinical illness who attend OPC | 0-5 yrs | 0.475 | 0.39 | 0.6 |  |
|  | 6-12 yrs | 0.118 | 0.09 | 0.17 |  |
|  | 13-17 yrs | 0.088 | 0.06 | 0.13 |  |
|  | 18-24 yrs | 0.035 | 0.02 | 0.08 |  |
|  | 25-44 yrs | 0.034 | 0.02 | 0.07 |  |
|  | 45-64 yrs | 0.027 | 0.01 | 0.05 |  |
|  | 65+ yrs | 0.036 | 0.02 | 0.07 |  |
| **DALY weights** | | | | | |
| DALY weights for influenza cases with mild illness/upper respiratory tract infection | All ages | 0.006 | 0.002 | 0.012 | (15) |
| DALY weights for influenza cases with lower respiratory tract illness that are not hospitalised | All ages | 0.051 | 0.032 | 0.074 |  |
| DALY weights for influenza cases with lower respiratory tract illness that are hospitalised | All ages | 0.133 | 0.088 | 0.19 |  |

*Table S5: Costs of influenza-associated illness (in USD) used in the economic model. Means and standard deviations (or means only where standard deviation values are not available) are presented. Samples of these parameters (except vaccine purchase costs) were drawn from a gamma distribution.*

| Type of cost | Mean | Standard deviation | Year | Reference |
| --- | --- | --- | --- | --- |
| **Indirect costs** | | | | |
| Child care costs among influenza cases attending outpatient clinics who report household members paid for child care | 0.07 | 0.57 | 2014 | (13) |
| Child care costs among hospitalised influenza cases who report household members paid for child care | 0.11 | 0.75 | 2014 |  |
| Lost wages among influenza cases attending outpatient visit who report that household members missed work | 12.84 | 27.17 | 2014 |  |
| Lost wages among hospitalised influenza cases who report that household members missed work | 42.02 | 41.54 | 2014 |  |
| **Direct medical costs** | | | | |
| Facility based medical costs among influenza cases attending outpatient clinic | 4.34 | 1.3 | 2014 | (13) |
| Facility based medical costs among hospitalised influenza cases | 59.19 | 59.39 | 2014 |  |
| Health care costs after discharge among hospitalised influenza cases who sought care after discharge | 3.28 | 6.19 | 2014 |  |
| Influenza vaccine purchase costs per dose | 3 | NA | - | Assumed |
| Vaccine administration cost per dose – provision of immunisation services at health facility | 1 | 0.72 | 2012 | (16) |
| Vaccine administration cost per dose - Supply chain cost per dose from national level to health facility | 0.43 | NA | 2012 |  |
| **Healthcare related costs** | | | | |
| Transportation costs among influenza cases with attending outpatient clinic | 0.4 | 0.87 | 2014 | (13) |
| Transportation costs among hospitalised influenza cases | 5.03 | 8.32 | 2014 |  |
| Cost of over the counter medication for influenza cases who had health care costs before visiting outpatient clinics | 1.39 | 3.9 | 2014 |  |
| Transportation costs to receive vaccine at health facility | 0.2 | 0.435 | - | Assumed |

*Table S6: Willingness to pay (WTP) thresholds used in this study. These include - the WHO ‘best buy’ threshold of $100 per DALY averted reserved for evaluating some of the most cost-effective programmes that WHO has ever evaluated* *(17)* *and thresholds derived using two broad approaches - marginal productivity thresholds which reflect opportunity costs of health interventions* *(18,19)**; and those based on values of national per capita gross domestic product (pc GDP) as estimated by the Commission for Macroeconomics and Health (CMH)* *(20,21)**.*

| Sl. No. | WTP threshold (USD) | Description* | Reference |
| --- | --- | --- | --- |
| 1. | 19.13 | 1% Kenya pc GDP (2019) | (19) |
| 2. | 100 | WHO best buy | (17) |
| 3. | 491 | 36% Kenya pc GDP (2015) | (18) |
| 4. | 497.29 | Median Kenya pc GDP (2019) (26% GDP) | (19) |
| 5. | 542 | 39% Kenya pc GDP (2015) | (18) |
| 6. | 623 | 45% Kenya pc GDP (2015) | (18) |
| 7. | 647 | 47% Kenya pc GDP (2015) | (18) |
| 8. | 975.45 | 51% Kenya pc GDP (2019) | (19) |
| 9. | 1912.65 | 1x Kenya pc GDP (2019) | (20,21) |
| 10. | 5737.95 | 3x Kenya pc GDP (2019) | (20,21) |

8.3 Calculation of threshold vaccine price

Threshold vaccine prices were calculated as

Threshold per-dose vaccine price = (Monetary benefit of vaccination)/(Number of vaccine doses administered)

where

Monetary benefit of vaccination = Total costs offset due to vaccination + (Total DALYs averted x willingness-to-pay per DALY averted)

8.4 Detailed results

*Table S7: Median values (and 95% credible intervals) for different measures for each of the six vaccination scenarios explored. Costs are expressed in terms of 2019 USD.*

| **Measure** | **No vaccination** | **Current seasonal** | **Improved (minimal)** | **Improved (breadth)** | **Improved (efficacy)** | **Universal** |
| --- | --- | --- | --- | --- | --- | --- |
| Total cases (millions) | 45.98 (38.72 53.45) | 32.41 (26.38 39.27) | 26.76 (20.81 34.07) | 22.14 (16.23 29.28) | 19.5 (13.86 26.51) | 15.39 (9.94 22.17) |
| Total URT infections (millions) | 26.88 (19.67 33.92) | 18.76 (13.87 24.96) | 15.51 (11.03 21.26) | 12.9 (8.77 18.02) | 11.34 (7.51 16.38) | 8.97 (5.62 13.71) |
| Total out-patient visits by patients with URT infection (millions) | 5.12 (3.95 6.48) | 3.25 (2.5 4.13) | 2.55 (1.92 3.25) | 2 (1.46 2.61) | 1.7 (1.22 2.28) | 1.27 (0.86 1.78) |
| Total LRT infections (millions) | 9.88 (6.18 14.4) | 6.96 (4.25 10.46) | 5.74 (3.42 8.97) | 4.75 (2.83 7.71) | 4.19 (2.45 6.9) | 3.28 (1.84 5.66) |
| Total LRT infections that are not hospitalised (millions) | 9.78 (6.11 14.3) | 6.9 (4.19 10.39) | 5.69 (3.38 8.91) | 4.71 (2.79 7.66) | 4.15 (2.42 6.84) | 3.25 (1.82 5.63) |
| Total patients with severe illness that are hospitalised | 0.1 (81868 0.13) | 68381 (53050 85874) | 54208 (41090 69508) | 43149 (30789 57266) | 36984 (25951 50111) | 28279 (18595 40494) |
| Total deaths | 9254 (4893 16746) | 6574 (3513 11830) | 5541 (2894 9904) | 4665 (2377 8575) | 4121 (2102 7624) | 3404 (1684 6433) |
| Total lost wages (millions USD) due to influenza related illness (non-hospitalised) | 7.96 (15 316.15) | 5.05 (10 209.7) | 3.86 (8 160.77) | 3.04 (6 126.68) | 2.59 (5 103.75) | 2.01 (4 80.41) |
| Total lost wages (millions USD) due to influenza related hospitalisation | 3.54 (0.12 18.81) | 2.33 (82826 12.39) | 1.83 (67894 9.72) | 1.49 (54155 7.98) | 1.26 (46350 6.7) | 0.97 (36148 5.27) |
| Total YLD (undiscounted) | 7290 (4194 11950) | 5164 (2912 8529) | 4281 (2320 7198) | 3542 (1851 6111) | 3091 (1601 5565) | 2459 (1201 4527) |
| Total YLL (undiscounted) (millions) | 0.31 (0.15 0.69) | 0.21 (0.1 0.46) | 0.17 (85391 0.36) | 0.14 (69106 0.29) | 0.12 (60488 0.25) | 96988 (46516 0.19) |
| Total DALYs (undiscounted) (millions) | 0.31 (0.16 0.69) | 0.21 (0.11 0.46) | 0.18 (87966 0.37) | 0.14 (72741 0.29) | 0.13 (62611 0.25) | 99551 (48614 0.2) |
| Total YLD (discounted) | 6462 (3710 10644) | 4585 (2555 7588) | 3813 (2075 6393) | 3152 (1652 5486) | 2759 (1429 4951) | 2202 (1086 4053) |
| Total YLL (discounted) (millions) | 0.15 (75763 0.3) | 0.1 (51355 0.2) | 85953 (43000 0.16) | 71190 (35767 0.13) | 62702 (31357 0.12) | 50639 (25062 97092) |
| Total DALYs (discounted) (millions) | 0.15 (81456 0.31) | 0.11 (55514 0.21) | 89982 (46239 0.17) | 74830 (38293 0.14) | 65722 (33293 0.12) | 52865 (26517 0.1) |
| Vaccine doses administered (including 15% wastage) (millions) | 0 | 34.98 | 34.98 | 16.14 | 22.42 | 16.14 |
| Vaccine costs (administration) (millions USD) | 0 | 62.96 (25.94 158.74) | 62.96 (25.94 158.74) | 30.45 (12.55 76.77) | 41.29 (17.01 104.09) | 30.45 (12.55 76.77) |
| Vaccine purchase costs (total doses) (millions USD) | 0 | 104.95 | 104.95 | 48.41 | 67.26 | 48.41 |
| Total vaccine costs (administration and purchase) (millions USD) | 0 | 167.91 (130.89 263.69) | 167.91 (130.89 263.69) | 78.86 (60.96 125.18) | 108.54 (84.27 171.35) | 78.86 (60.96 125.18) |
| Direct medical costs (millions USD) | 38.22 (19.29 70.88) | 192.45 (152.43 293.68) | 187.05 (147.41 285.48) | 94.58 (73.54 143.04) | 121.85 (95.14 186.48) | 89.15 (69.57 136.77) |
| Direct medical costs (excl. vaccine purchase and administration costs) (millions USD) | 38.22 (19.29 70.88) | 24.6 (12.12 45.91) | 19.19 (9.18 35.82) | 15.42 (7.34 29.12) | 13.09 (6.24 25.13) | 10.07 (4.59 19.64) |
| Healthcare related costs (millions USD) | 3.96 (0.03 68.04) | 6.37 (0.03 84.23) | 5.59 (0.02 74.93) | 3.5 (0.02 45.19) | 3.75 (0.02 49.17) | 2.86 (0.01 36.66) |
| Healthcare related costs (excl. transport costs to seek vaccination) (millions USD) | 3.96 (0.03 68.04) | 2.57 (0.02 43.61) | 2.02 (0.02 34.47) | 1.6 (0.01 26.43) | 1.36 (0.01 22.34) | 1.05 (0.01 16.67) |
| Indirect costs (millions USD) | 14.79 (0.74 319.01) | 9.59 (0.48 215.64) | 7.58 (0.38 168.18) | 6.01 (0.29 132.29) | 5.08 (0.25 106.95) | 4 (0.2 80.83) |
| Total costs (millions USD) | 69.7 (29.63 392.52) | 230.73 (165.56 463.21) | 220.56 (159.27 403.95) | 115.89 (81.2 253.7) | 143.54 (103.34 271.65) | 105.38 (75.76 198.59) |
| Total costs (excl. vaccine purchase administration and transport costs) (millions USD) | 69.7 (29.63 392.52) | 44.87 (19.15 250.77) | 35.31 (14.87 192.27) | 28.62 (12.04 151.51) | 24.28 (10.06 129.82) | 18.68 (7.43 99.23) |
| Discounted total costs (millions USD) | 62.68 (26.45 351.54) | 206.33 (147.52 415.91) | 196.97 (141.89 362.76) | 105.19 (73.46 232.49) | 129.15 (92.78 245.28) | 95.65 (68.65 182.97) |
| Incremental total costs (millions USD) | 0 | 144.02 (40.55 255.28) | 135.01 (-14.59 246.96) | 39.34 (-140.89 91.66) | 64.54 (-134.41 137.06) | 29.81 (-197.52 85.18) |
| Incremental total costs (discounted) (millions USD) | 0 | 128.64 (35.62 228.43) | 120.28 (-15.77 221.4) | 36.21 (-123.05 84.66) | 58.17 (-118.79 123.59) | 27.67 (-174.38 78.21) |
| DALYs averted (undiscounted) (millions) | 0 | 0.1 (0.04 0.25) | 0.14 (0.06 0.34) | 0.17 (0.08 0.41) | 0.19 (0.09 0.45) | 0.22 (0.1 0.5) |
| ICER per DALY averted (discounted costs) | NA | 1266 (257 3601) | 833 (-79 2415) | 194 (-711 722) | 289 (-605 950) | 122 (-821 511) |
| DALYs averted (discounted) (millions) | 0 | 0.05 (0.02 0.1) | 0.06 (0.03 0.14) | 0.08 (0.04 0.17) | 0.09 (0.04 0.19) | 0.1 (0.05 0.21) |
| ICER per DALY averted (discounted costs & DALYs) | NA | 2764 (635 7494) | 1807 (-176 4772) | 422 (-1548 1453) | 626 (-1317 1910) | 263 (-1698 1061) |

*Table S8: Total vaccine doses administered and median (with 95% credible intervals) values of total cases total discounted DALYs incurred total undiscounted costs and total discounted DALYs averted in each year for each vaccination scenario. All costs are expressed in terms of 2019 USD.*

| **Vaccine** | **Year** | **Total doses administered (millions)** | **Total cases (millions)** | **Total DALYs (discounted)** | **Total costs (discounted) (millions USD)** | **Total DALYs averted (discounted)** |
| --- | --- | --- | --- | --- | --- | --- |
| No vaccination | 2010 | 0 | 2.66 (0.88 5) | 10814 (3214 27198) | 5.45 (1.28 33.84) | 0 |
|  | 2011 |  | 6.82 (4.51 9.59) | 23272 (10591 48613) | 11.54 (4.61 63.95) | 0 |
|  | 2012 |  | 7.44 (5.1 10.63) | 30781 (13473 64102) | 12.76 (5.24 68.26) | 0 |
|  | 2013 |  | 2.29 (0.63 4.17) | 7101 (2195 21715) | 4.05 (0.8 23.9) | 0 |
|  | 2014 |  | 4.62 (2.86 6.32) | 12990 (5367 28689) | 5.37 (2.07 30.06) | 0 |
|  | 2015 |  | 0.65 (0.52 0.82) | 2535 (1350 4591) | 0.72 (0.32 3.98) | 0 |
|  | 2016 |  | 14.87 (11.12 19.91) | 44930 (20222 91797) | 14.41 (5.9 76.07) | 0 |
|  | 2017 |  | 0.6 (0.42 1.27) | 2446 (1125 6510) | 0.72 (0.25 4.48) | 0 |
|  | 2018 |  | 5.5 (3.17 8.08) | 16582 (6252 44415) | 6.78 (2.53 37.14) | 0 |
|  | 2019 |  | 0.25 (0.15 0.38) | 774 (330 1953) | 0.28 (97506 1.53) | 0 |
| Current seasonal | 2010 | 3.49 | 0.91 (0.5 2.77) | 4204 (1737 12745) | 22.96 (16.16 40.36) | 6303 (1275 16514) |
|  | 2011 | 3.58 | 4.83 (2.94 7.42) | 16376 (6959 34047) | 28.44 (19.48 66.43) | 6708 (2942 16110) |
|  | 2012 | 3.68 | 6.47 (4.14 9.88) | 26391 (11242 55663) | 30.27 (20.2 77.66) | 4129 (1555 8919) |
|  | 2013 | 3.78 | 0.62 (0.42 1.22) | 2437 (1239 5649) | 19.29 (14.32 31.86) | 4733 (639 16427) |
|  | 2014 | 3.88 | 4.03 (2.14 5.75) | 11014 (4332 25066) | 22.63 (16.25 45.41) | 1831 (589 4288) |
|  | 2015 | 3.98 | 0.64 (0.51 0.83) | 2479 (1315 4403) | 17.63 (13.54 27.5) | 47 (-130 226) |
|  | 2016 | 4.09 | 11.89 (8.17 17.48) | 35682 (15488 71319) | 28.11 (18.88 76.32) | 9426 (3421 22424) |
|  | 2017 | 4.19 | 0.45 (0.38 0.69) | 1889 (989 3709) | 16.45 (12.95 25.07) | 517 (86 3077) |
|  | 2018 | 4.3 | 1.72 (0.92 3.36) | 5234 (2181 14176) | 17.95 (13.78 29.58) | 11213 (4011 30291) |
|  | 2019 | 0 | 0.18 (70168 0.39) | 540 (192 1780) | 0.19 (42836 1.25) | 208 (-671 1243) |
| Improved (minimal) | 2010 | 3.49 | 0.89 (0.49 2.76) | 4146 (1713 12606) | 22.9 (16.13 40.32) | 6347 (1313 16626) |
|  | 2011 | 3.58 | 3.73 (2.15 6.13) | 12633 (5146 25531) | 26.18 (18.08 55.35) | 10356 (4601 24796) |
|  | 2012 | 3.68 | 5.26 (2.98 8.79) | 21212 (8451 47379) | 27.94 (18.9 66.65) | 9205 (3959 19522) |
|  | 2013 | 3.78 | 0.53 (0.4 0.88) | 2174 (1153 4439) | 19.01 (14.18 31.02) | 5007 (689 17931) |
|  | 2014 | 3.88 | 3.09 (1.03 4.87) | 8108 (2775 20229) | 21.34 (15.55 37.79) | 4622 (1461 11093) |
|  | 2015 | 3.98 | 0.69 (0.53 0.91) | 2592 (1357 4700) | 17.71 (13.55 27.76) | -34 (-692 380) |
|  | 2016 | 4.09 | 10.28 (6.91 16.23) | 30438 (13151 62850) | 26.4 (17.95 69.03) | 14133 (5447 33053) |
|  | 2017 | 4.19 | 0.41 (0.37 0.55) | 1735 (945 3107) | 16.33 (12.89 24.9) | 654 (115 3724) |
|  | 2018 | 4.3 | 1.09 (0.68 2.31) | 3528 (1601 9124) | 17.02 (13.3 25.98) | 13014 (4527 35846) |
|  | 2019 | 0 | 0.1 (64653 0.34) | 377 (167 1317) | 0.11 (31224 0.92) | 376 (-427 1422) |
| Improved (breadth) | 2010 | 3.49 | 0.88 (0.48 2.74) | 4107 (1666 12484) | 22.81 (16.1 40.27) | 6386 (1344 16748) |
|  | 2011 | 1.44 | 3.34 (1.81 5.7) | 11242 (4628 23413) | 13.04 (8.48 34.55) | 11597 (5207 27766) |
|  | 2012 | 1.48 | 4.72 (2.54 8.34) | 18798 (7191 43569) | 15.11 (9.34 48.78) | 11456 (4962 23921) |
|  | 2013 | 1.52 | 0.58 (0.4 1.11) | 2298 (1187 5167) | 8.39 (6.1 14.23) | 4874 (705 17154) |
|  | 2014 | 1.56 | 1.9 (0.51 3.93) | 5023 (1615 15761) | 9.31 (6.43 19.04) | 7538 (2445 17926) |
|  | 2015 | 1.6 | 0.71 (0.51 0.99) | 2592 (1327 4831) | 7.65 (5.79 12.34) | -40 (-1038 719) |
|  | 2016 | 1.64 | 8.07 (4.44 14.33) | 23239 (9365 50966) | 13.68 (8.45 45.32) | 21019 (8283 46493) |
|  | 2017 | 1.68 | 0.4 (0.37 0.52) | 1692 (917 2996) | 6.86 (5.34 10.46) | 722 (138 3873) |
|  | 2018 | 1.73 | 0.64 (0.51 1.02) | 2274 (1172 4822) | 7.04 (5.47 11.21) | 14271 (4902 39024) |
|  | 2019 | 0 | 64814 (62121 0.12) | 268 (144 497) | 56614 (23857 0.35) | 516 (134 1535) |
| Improved (efficacy) | 2010 | 3.49 | 0.75 (0.46 2.36) | 3485 (1542 10281) | 22.48 (15.82 39) | 6994 (1454 18214) |
|  | 2011 | 2.15 | 2.79 (1.44 5.05) | 9261 (3775 19929) | 16.08 (10.99 34.21) | 13329 (6097 31484) |
|  | 2012 | 2.21 | 3.97 (1.91 7.88) | 15766 (5690 38873) | 17.9 (11.67 44.96) | 14279 (6143 30017) |
|  | 2013 | 2.27 | 0.48 (0.39 0.72) | 2011 (1086 3816) | 11.74 (8.67 19.2) | 5188 (737 18469) |
|  | 2014 | 2.33 | 1.48 (0.44 3.63) | 4080 (1420 14222) | 12.42 (9.01 21.61) | 8306 (2779 20110) |
|  | 2015 | 2.39 | 0.7 (0.47 0.99) | 2507 (1295 4720) | 10.97 (8.43 17.14) | 8 (-1035 826) |
|  | 2016 | 2.46 | 7.46 (3.78 13.56) | 21452 (8114 47692) | 16.34 (10.98 45.54) | 22839 (9017 52614) |
|  | 2017 | 2.52 | 0.39 (0.36 0.48) | 1638 (909 2820) | 9.99 (7.84 15.18) | 769 (145 4053) |
|  | 2018 | 2.58 | 0.58 (0.49 0.86) | 2107 (1112 4157) | 10.07 (7.94 15.31) | 14388 (4960 39431) |
|  | 2019 | 0 | 62856 (61873 91319) | 258 (143 440) | 52442 (23252 0.32) | 529 (155 1560) |
| Universal | 2010 | 3.49 | 0.74 (0.45 2.36) | 3447 (1523 10245) | 22.46 (15.8 38.91) | 7027 (1469 18255) |
|  | 2011 | 1.44 | 2.52 (1.29 4.71) | 8423 (3316 18448) | 11.59 (7.75 27.13) | 14155 (6488 33798) |
|  | 2012 | 1.48 | 3.23 (1.43 7.34) | 12680 (4369 34604) | 12.7 (7.99 34.62) | 17147 (7528 36174) |
|  | 2013 | 1.52 | 0.47 (0.38 0.67) | 1934 (1044 3593) | 8.04 (5.92 13.12) | 5259 (743 18659) |
|  | 2014 | 1.56 | 0.71 (0.37 2.84) | 2438 (1155 9429) | 8.11 (5.96 14.05) | 10007 (3624 22838) |
|  | 2015 | 1.6 | 0.57 (0.41 0.98) | 2213 (1171 4290) | 7.43 (5.69 11.66) | 334 (-835 1214) |
|  | 2016 | 1.64 | 5.53 (1.17 10.78) | 15177 (3320 35786) | 10.91 (7.09 30.86) | 29101 (12154 65478) |
|  | 2017 | 1.68 | 0.37 (0.35 0.51) | 1597 (884 2816) | 6.8 (5.3 10.37) | 815 (118 4178) |
|  | 2018 | 1.73 | 0.44 (0.4 0.54) | 1679 (936 3006) | 6.73 (5.28 10.21) | 14780 (5115 41184) |
|  | 2019 | 0 | 59676 (59673 59958) | 239 (133 395) | 44610 (20062 0.24) | 547 (173 1593) |

*Table S9: Median (and 95% CrI) values of incremental net monetary benefits (in millions of USD) (2010 - 2019) across ten thresholds of willingness-to-pay per DALY averted.*

| Vaccine | Threshold = $19 | Threshold = $100 | Threshold = $491 | Threshold = $497 | Threshold = $542 | Threshold = $623 | Threshold = $647 | Threshold = $975 | Threshold = $1913 | Threshold = $5738 |
| --- | --- | --- | --- | --- | --- | --- | --- | --- | --- | --- |
| Current seasonal | -127.76 (-227.59, -34.25) | -123.6 (-223.98, -28.43) | -104.32 (-202.27, -9.2) | -103.95 (-201.98, -8.84) | -101.78 (-199.92, -6.25) | -97.93 (-195.24, -1.16) | -96.93 (-194.5, 1.53) | -82.25 (-183.08, 31.48) | -39.2 (-143.13, 109.39) | 134.63 (-39.09, 489.8) |
| Improved (minimal) | -118.8 (-220.36, 17.39) | -113.16 (-214.87, 21.92) | -86.66 (-186.19, 49.73) | -86.33 (-185.91, 50.23) | -82.91 (-183.92, 53.8) | -77.25 (-180.14, 60.26) | -75.62 (-178.75, 62.18) | -54.74 (-157.99, 98.24) | 6.99 (-117.26, 213.75) | 255.81 (30.19, 718.63) |
| Improved (breadth) | -34.53 (-83.51, 124.32) | -27.78 (-76.78, 131.38) | 6.07 (-50.61, 169.52) | 6.61 (-50.29, 170.15) | 10.25 (-47.46, 174.6) | 17.26 (-43.14, 182.62) | 19.24 (-41.88, 185) | 46.62 (-24.81, 222.25) | 124.21 (21.71, 366.2) | 431.99 (179.62, 989.82) |
| Improved (efficacy) | -56.66 (-122.62, 120.29) | -49.75 (-114.27, 127.88) | -12.73 (-85.26, 170.01) | -12.02 (-84.85, 170.7) | -7.66 (-81.75, 175.56) | -0.28 (-76.04, 184.34) | 2.03 (-74.25, 186.94) | 32.85 (-52.92, 226.31) | 118.95 (0.18, 385.89) | 462.42 (181.64, 1073.65) |
| Universal | -25.75 (-76.31, 176.39) | -17.89 (-68.04, 185.56) | 24.7 (-37.31, 242.23) | 25.42 (-36.81, 242.88) | 30.52 (-33.22, 247.46) | 39.6 (-28.45, 256.07) | 41.92 (-26.46, 258.66) | 77.79 (-4.39, 308.53) | 176.96 (52.96, 467.51) | 566.33 (258.78, 1232.09) |

*Table S10: Median (and 95% CrI) values of threshold per-dose vaccine prices at or below which each vaccination scenario is cost-effective calculated using discounted costs and DALYs across ten thresholds of willingness-to-pay per DALY averted. These are calculated while including a median vaccine administration cost of approximately $1.80 per dose.*

| Vaccine | Threshold = $19 | Threshold = $100 | Threshold = $491 | Threshold = $497 | Threshold = $542 | Threshold = $623 | Threshold = $647 | Threshold = $975 | Threshold = $1913 | Threshold = $5738 |
| --- | --- | --- | --- | --- | --- | --- | --- | --- | --- | --- |
| Current seasonal | -0.99 (-3.85, 1.68) | -0.87 (-3.74, 1.85) | -0.32 (-3.12, 2.4) | -0.31 (-3.11, 2.41) | -0.25 (-3.06, 2.48) | -0.14 (-2.92, 2.63) | -0.11 (-2.9, 2.7) | 0.31 (-2.57, 3.56) | 1.54 (-1.43, 5.79) | 6.51 (1.54, 16.66) |
| Improved (minimal) | -0.74 (-3.64, 3.16) | -0.58 (-3.48, 3.29) | 0.18 (-2.66, 4.08) | 0.19 (-2.65, 4.1) | 0.29 (-2.6, 4.2) | 0.45 (-2.49, 4.38) | 0.5 (-2.45, 4.44) | 1.09 (-1.86, 5.47) | 2.86 (-0.69, 8.77) | 9.97 (3.52, 23.2) |
| Improved (breadth) | 0.56 (-2.47, 10.41) | 0.98 (-2.05, 10.85) | 3.08 (-0.43, 13.21) | 3.11 (-0.41, 13.25) | 3.34 (-0.24, 13.52) | 3.77 (0.03, 14.02) | 3.9 (0.11, 14.17) | 5.59 (1.17, 16.48) | 10.4 (4.05, 25.4) | 29.47 (13.83, 64.04) |
| Improved (efficacy) | 0.15 (-2.79, 8.05) | 0.46 (-2.42, 8.38) | 2.11 (-1.12, 10.26) | 2.14 (-1.1, 10.29) | 2.34 (-0.97, 10.51) | 2.67 (-0.71, 10.9) | 2.77 (-0.63, 11.02) | 4.15 (0.32, 12.78) | 7.99 (2.69, 19.89) | 23.31 (10.78, 50.57) |
| Universal | 1.11 (-2.03, 13.63) | 1.59 (-1.51, 14.2) | 4.23 (0.39, 17.71) | 4.28 (0.42, 17.75) | 4.6 (0.64, 18.04) | 5.16 (0.94, 18.57) | 5.3 (1.06, 18.73) | 7.52 (2.43, 21.82) | 13.67 (5.99, 31.68) | 37.8 (18.74, 79.06) |

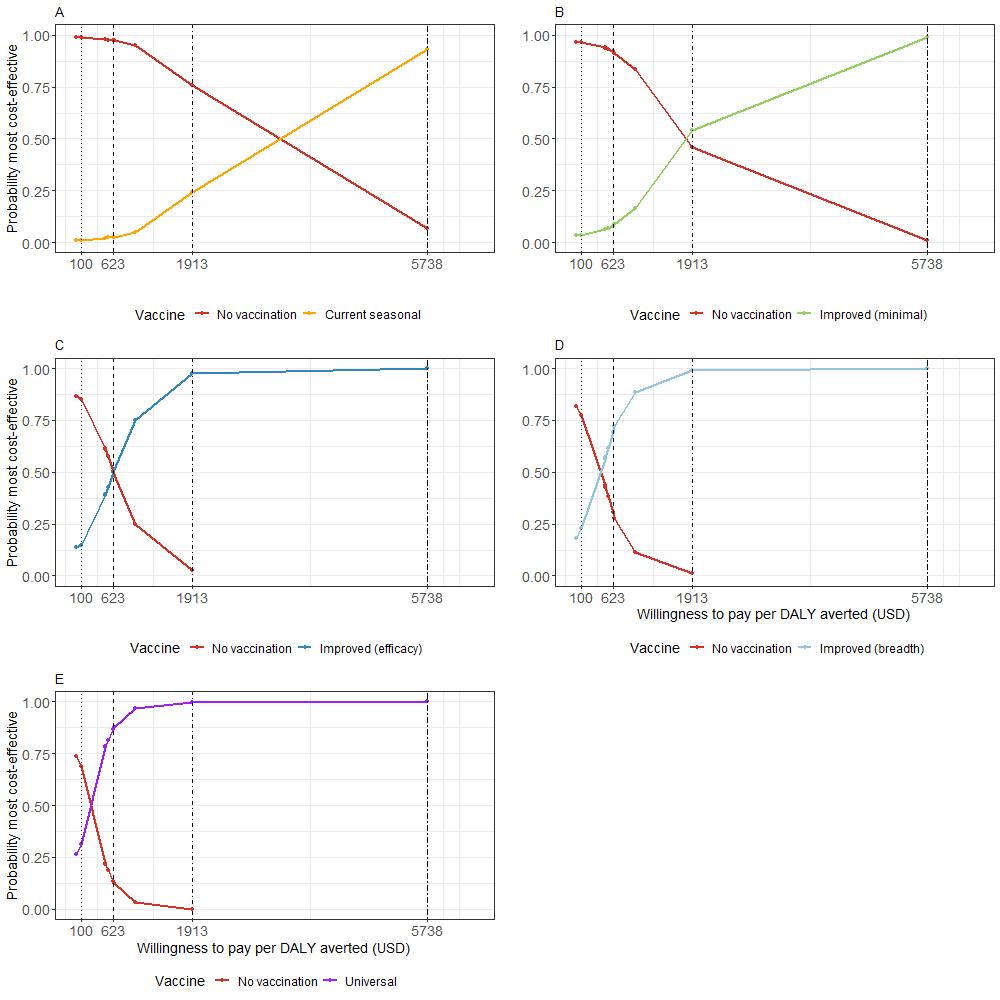

*Figure S7: Cost effectiveness acceptability curves for the five different vaccine scenarios. Vertical lines represent four selected willingness to pay thresholds (USD) per DALY averted. A. Current seasonal vaccine B. Improved (minimal) vaccine C. Improved (efficacy) vaccine D. Improved (breadth) vaccine D. Universal vaccine*

### 9. Sensitivity Analysis

A. Coverage

A sensitivity analysis was run with increased coverage of vaccines in the target age groups. This coverage was increased to 75% but all the other aspects of the modelling were kept the same as in the base case. Figures S8-S9 show the epidemiological model output and Figure S10 shows the cost-effectiveness output.

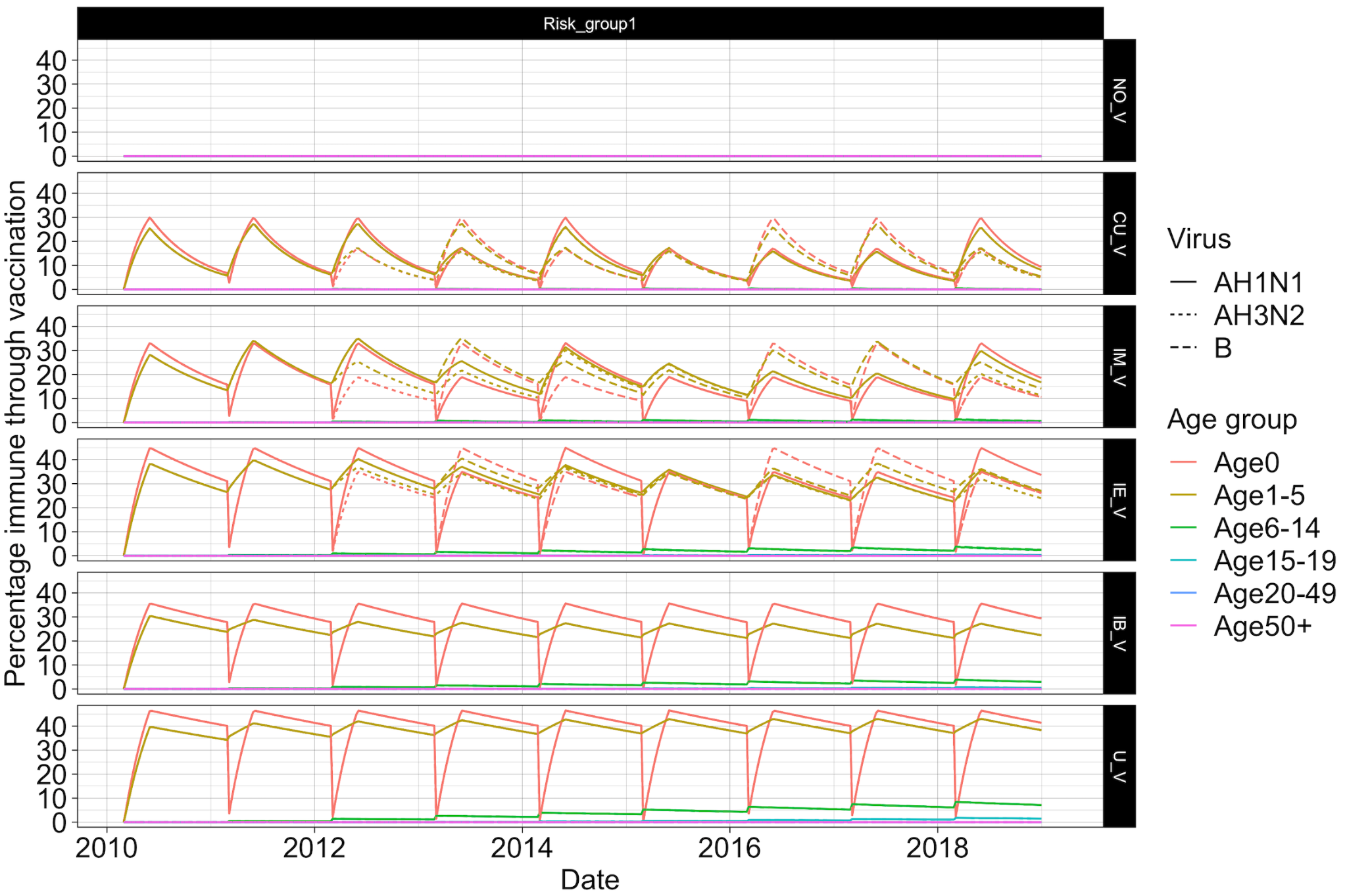

*Figure S8: Percentage of the population with vaccine immunity for each scenario split by virus subtype and age group for a sensitivity analysis with 75% coverage across target age groups as opposed to the main analysis scenario of 50%.*

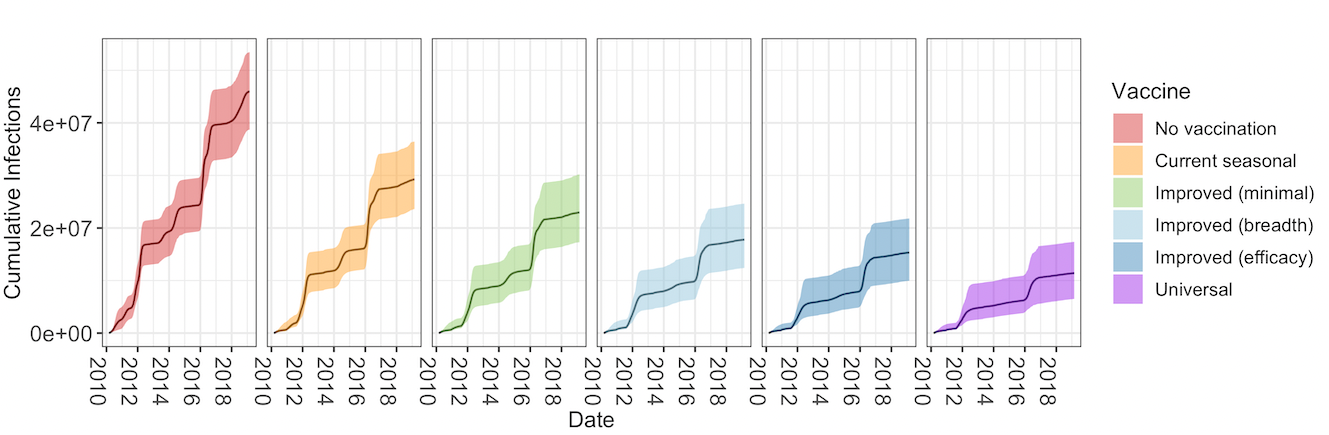

*Figure S9: Cumulative number of infections by vaccine scenario when vaccine coverage is 75%.*

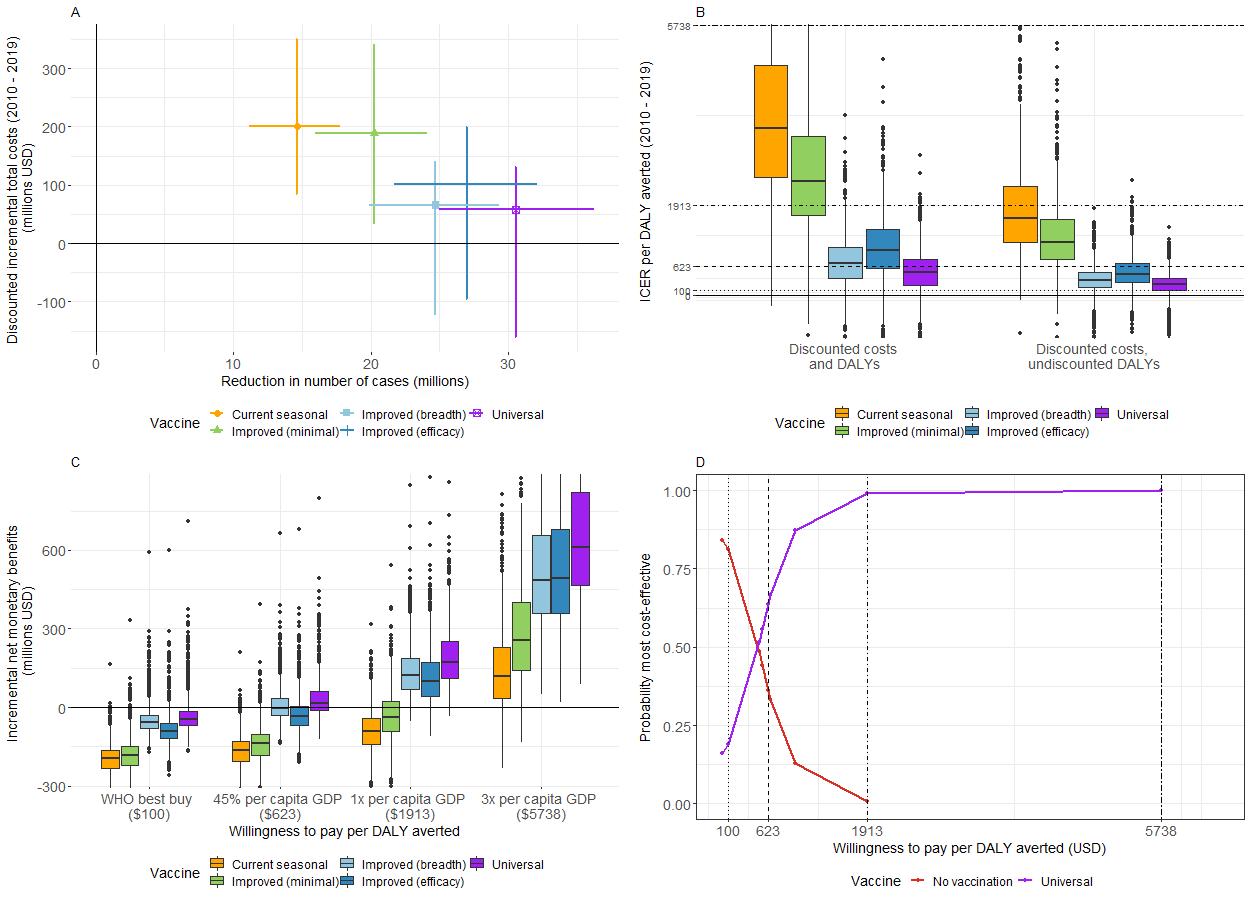

*Figure S10: Results with vaccine coverage at 75%. A) Mean (with 95% CrI) reduction in number of cases (in millions) by mean (with 95% CrI) discounted incremental total costs (in millions of USD) for each vaccine (2010 – 2019). B) Boxplot of incremental cost effectiveness ratios (ICER) per disability adjusted life year (DALY) averted for each vaccine (2010 - 2019). Horizontal lines represent different willingness-to-pay thresholds per DALY averted. C) Boxplot of incremental net monetary benefits (INMB) (in millions of USD) (2010 to 2019) at four selected thresholds of willingness-to-pay per DALY averted. D) Cost-effectiveness acceptability curve for vaccination scenarios with the highest probabilities of being most cost-effective at four selected willingness-to-pay thresholds per DALY averted (vertical lines).*

B. Susceptibility - population fraction with waning.

In this sensitivity analysis we assumed that lack of infections as a result of vaccinations resulted in an increased susceptibility to the same subtype (or lineage in the case of flu B) in the following years. We increased the susceptibility of the population by the fraction of the population that averted infection by the same subtype in all previous years reduced by an annual percentage of 10% to account for waning of infection-derived immunity in the original model. Modelling results are shown in Figure S11 where across all vaccine scenarios vaccination resulted in more infections. The relevant cost-effectiveness analysis results are shown in Figure S12.

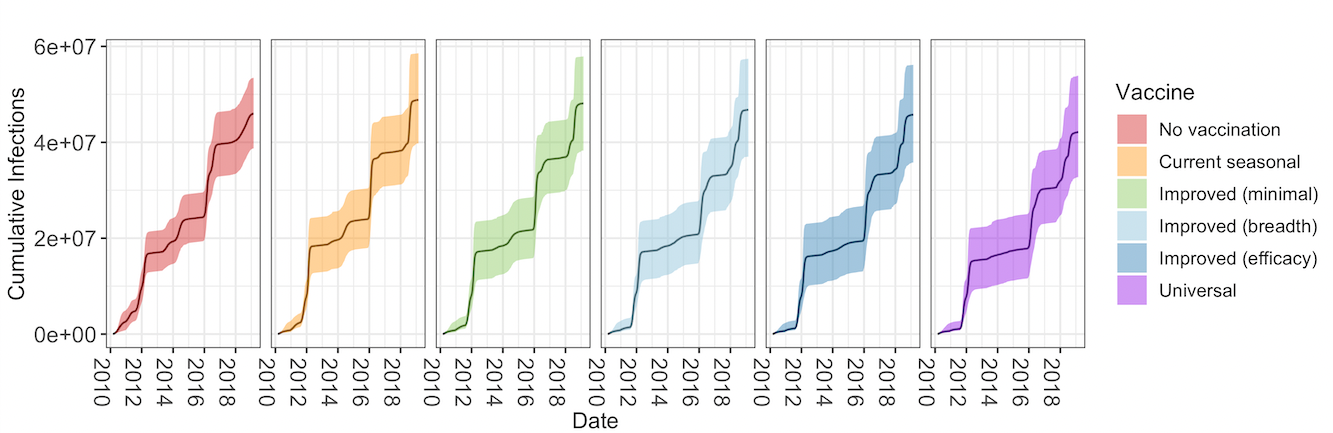
*Figure S11: Scenario with increased susceptibility after vaccination : Cumulative number of infections by vaccine scenario.*

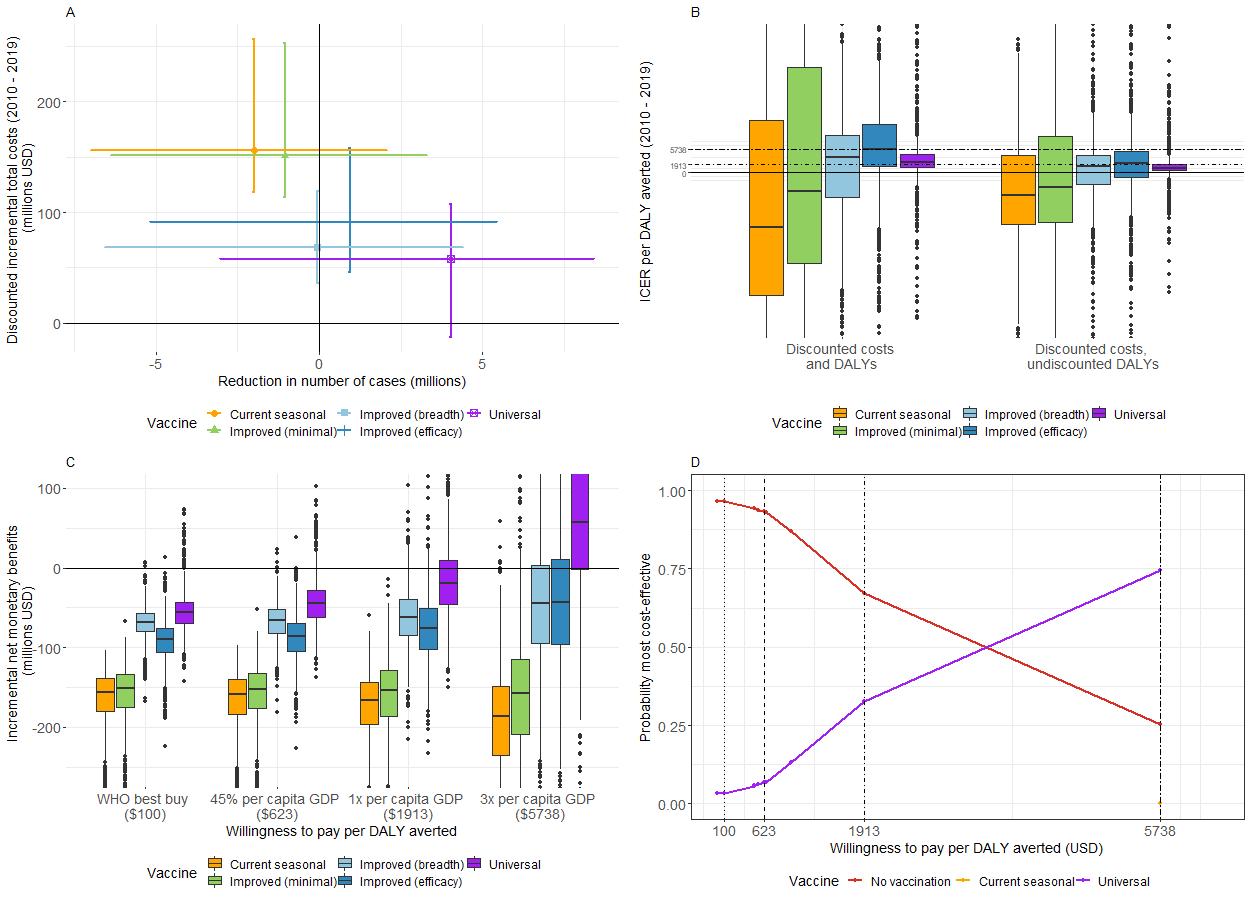

*Figure S12: Scenario with increased susceptibility after vaccination. A) Mean (with 95% CrI) reduction in number of cases (in millions) by mean (with 95% CrI) discounted incremental total costs (in millions of USD) for each vaccine (2010 – 2019). B) Boxplot of incremental cost effectiveness ratios (ICER) per disability adjusted life year (DALY) averted for each vaccine (2010 - 2019). Horizontal lines represent different willingness-to-pay thresholds per DALY averted. C) Boxplot of incremental net monetary benefits (INMB) (in millions of USD) (2010 to 2019) at four selected thresholds of willingness-to-pay per DALY averted. D) Cost-effectiveness acceptability curve for vaccination scenarios with the highest probabilities of being most cost-effective at four selected willingness-to-pay thresholds per DALY averted (vertical lines).*

C. Susceptibility - fixed reduction

In this sensitivity analysis we also assumed that a lack of infection as a result of vaccination resulted in a reduction in natural immunity (i.e. increased susceptibility). Here we assumed a constant 20% increase in susceptibility in ages 0-5	across all seasons where vaccination had been implemented in the season before. Figure S13 shows the epidemiological and Figure S14 shows the cost-effectiveness analysis.

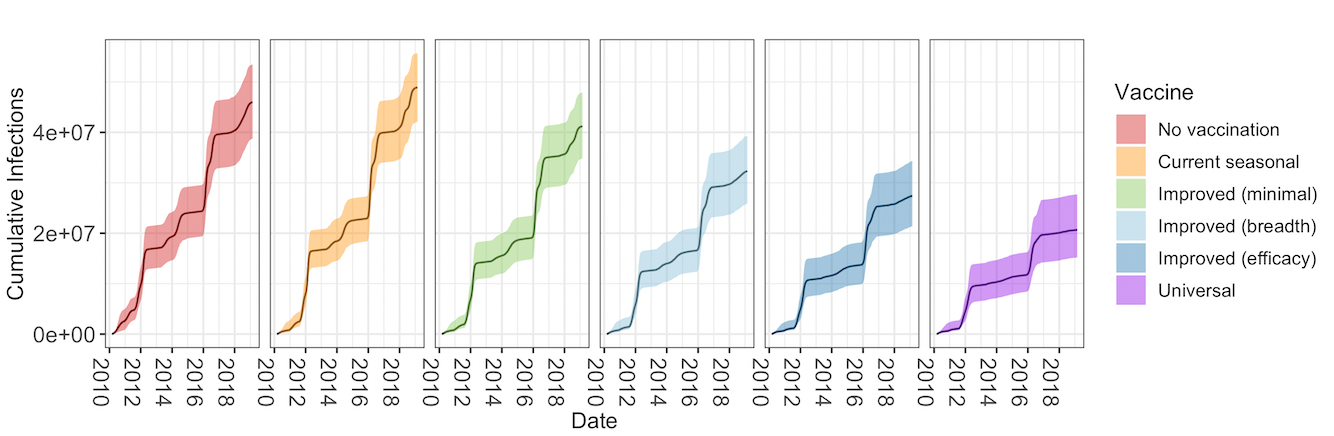

*Figure S13: Sensitivity analysis with a fixed reduction in susceptibility: Cumulative number of infections by vaccine scenario.*

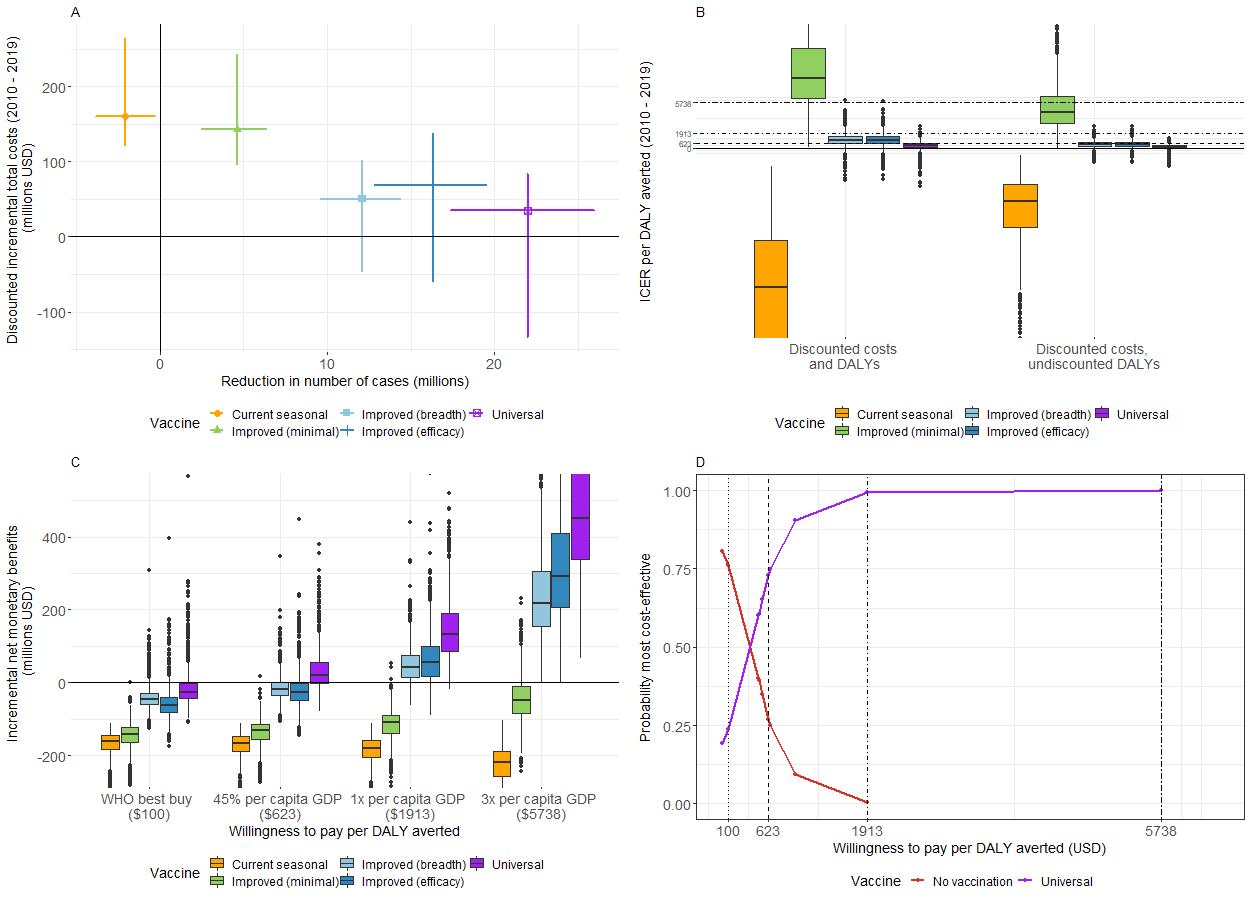

*Figure S14: Sensitivity analysis with a fixed reduction in susceptibility: A) Mean (with 95% CrI) reduction in number of cases (in millions) by mean (with 95% CrI) discounted incremental total costs (in millions of USD) for each vaccine (2010 – 2019). B) Boxplot of incremental cost effectiveness ratios (ICER) per disability adjusted life year (DALY) averted for each vaccine (2010 - 2019). Horizontal lines represent different willingness-to-pay thresholds per DALY averted. C) Boxplot of incremental net monetary benefits (INMB) (in millions of USD) (2010 to 2019) at four selected thresholds of willingness-to-pay per DALY averted. D) Cost-effectiveness acceptability curve for vaccination scenarios with the highest probabilities of being most cost-effective at four selected willingness-to-pay thresholds per DALY averted (vertical lines).*

D. Exact efficacies

We ran the model using the efficacies for current seasonal and improved (minimal) vaccines found in the literature as in table S1, as opposed to the assumed of 40% or 70% efficacy dependent on match or mismatch.

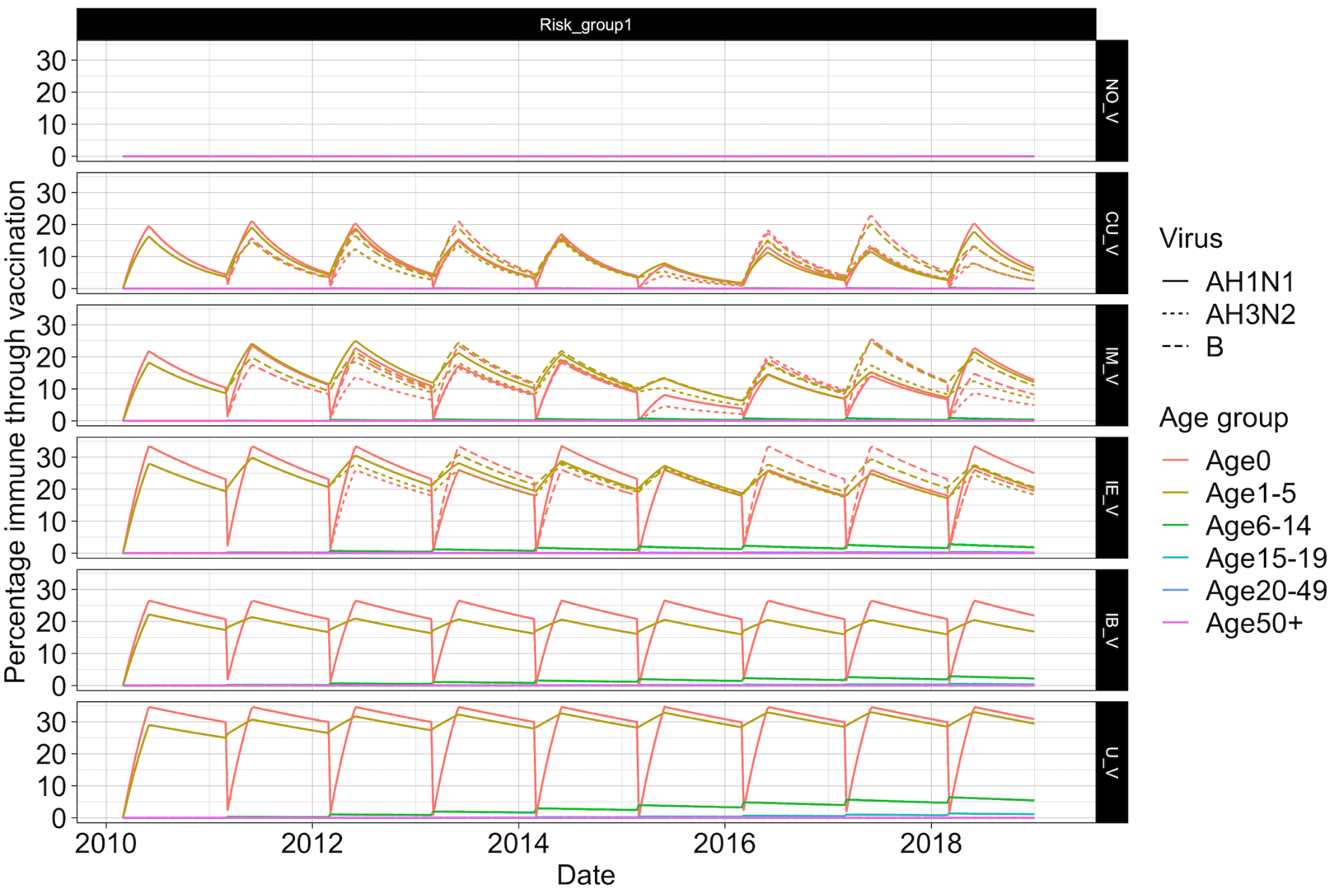

*Figure S15: Exact efficacies sensitivity analysis: Percentage of the population with vaccine immunity for each scenario split by virus subtype and age group*

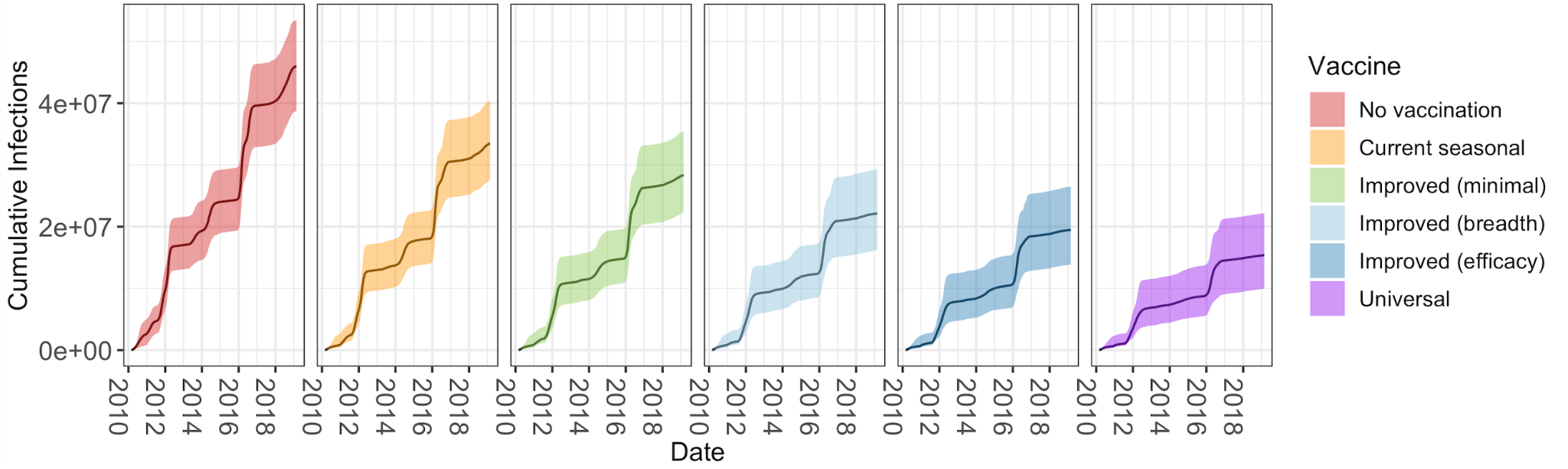

*Figure S16: Exact efficacies sensitivity analysis: Cumulative number of infections by vaccine scenario.*

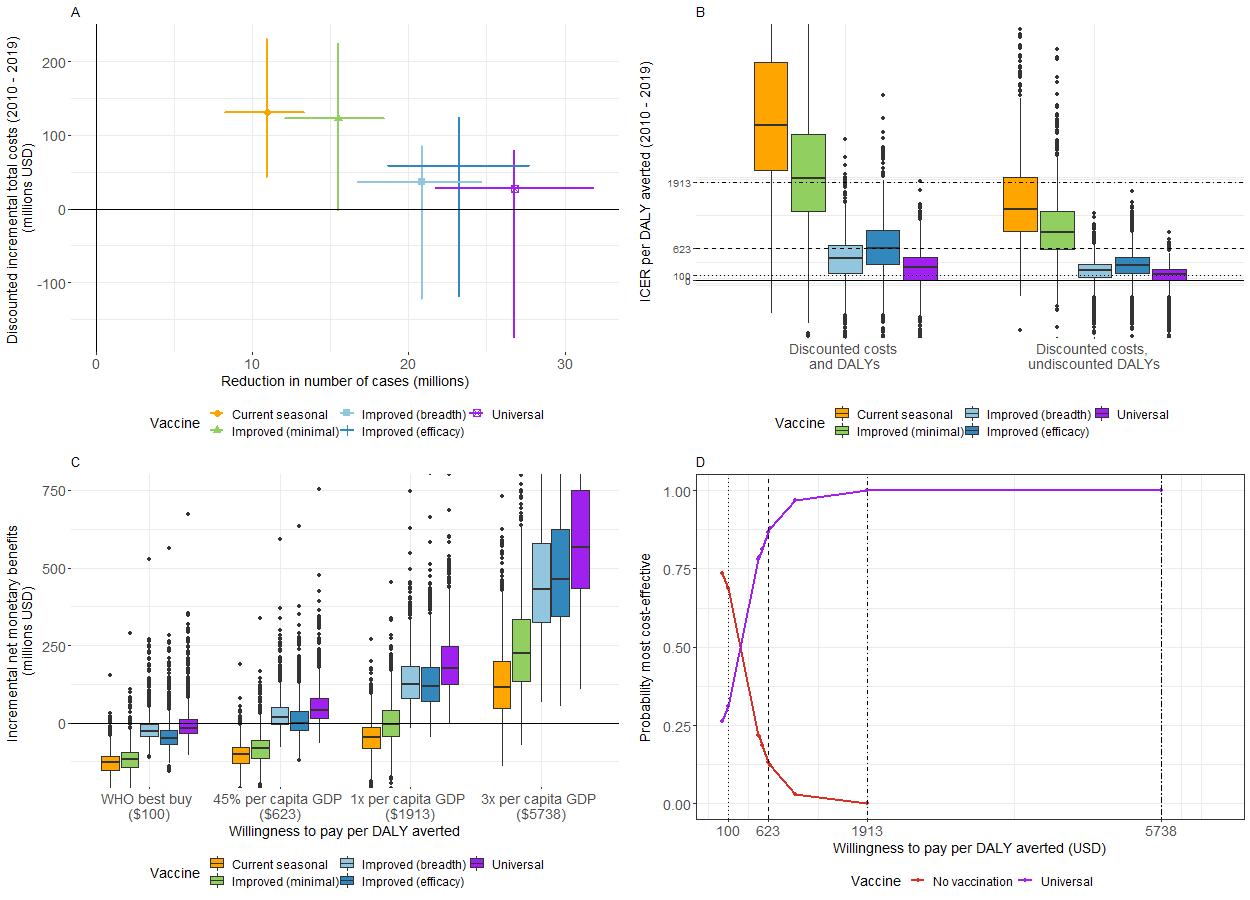

*Figure S17: Exact efficacies sensitivity analysis: A) Mean (with 95% CrI) reduction in number of cases (in millions) by mean (with 95% CrI) discounted incremental total costs (in millions of USD) for each vaccine (2010 – 2019). B) Boxplot of incremental cost effectiveness ratios (ICER) per disability adjusted life year (DALY) averted for each vaccine (2010 - 2019). Horizontal lines represent different willingness-to-pay thresholds per DALY averted. C) Boxplot of incremental net monetary benefits (INMB) (in millions of USD) (2010 to 2019) at four selected thresholds of willingness-to-pay per DALY averted. D) Cost-effectiveness acceptability curve for vaccination scenarios with the highest probabilities of being most cost-effective at four selected willingness-to-pay thresholds per DALY averted (vertical lines).*

E. Vaccine price

We ran an additional sensitivity analysis altering the cost per vaccine dose to $1.5, $6 or $10. Cost-effectiveness acceptability curves are shown in Figure S18. As per dose vaccine price increases the universal vaccines are cost-effective compared to no vaccination only at higher WTP thresholds.

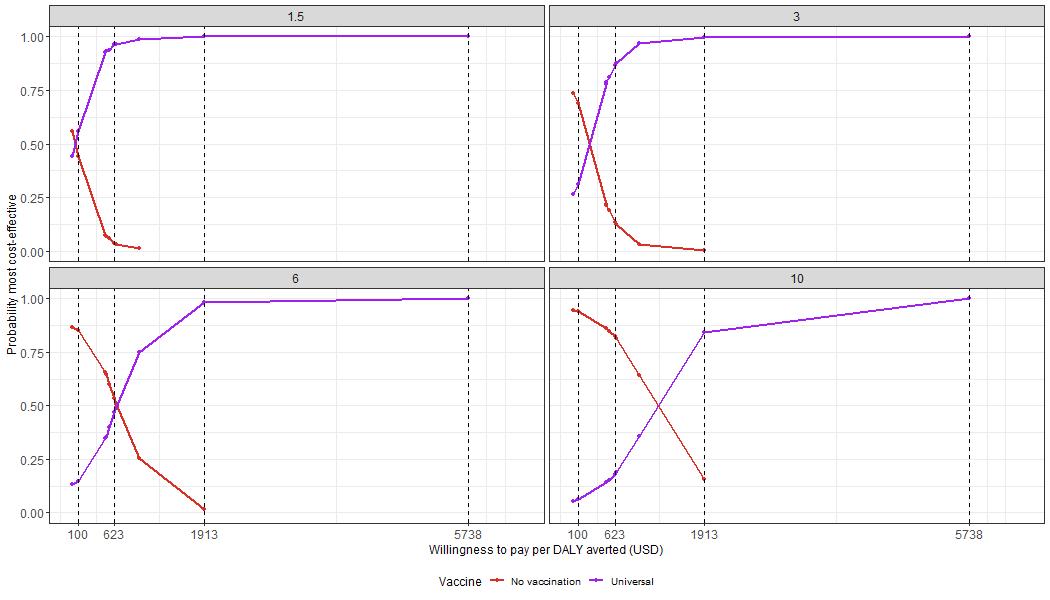

*Figure S18: Cost-effectiveness acceptability curves for the universal vaccination scenario with the highest probabilities of being most cost-effective at four selected willingness-to-pay thresholds per DALY averted (vertical lines) and assuming four different per-dose vaccine prices ($1.5 $3 $6 and $10)*
